# ECG-based longitudinal risk prediction across diseases and organ systems

**DOI:** 10.64898/2026.08.29.26361697

**Authors:** Yilong Ye, Zhiwei Zeng, Xin Tian, Zhenming Yuan, Jinghua Wang, Yajie Zhu

## Abstract

Artificial intelligence applied to routine electrocardiograms (ECGs) has largely focused on detecting existing disease or predicting individual cardiovascular outcomes. Whether ECGs can support prediction of multiple future diseases across organ systems remains unclear. We developed ECG-RISK, a multitask survival model for 67 incident three-character ICD-10 endpoints using ECG waveforms, demographic characteristics and routinely collected laboratory data from 86,673 MIMIC-IV patients. Discrimination was highest for heart, brain, kidney and lung endpoints, with organ-level C-indices ranging from 0.796 to 0.825, whereas liver and pancreatic endpoints showed lower discrimination. The ECG-only model achieved strong discrimination across most endpoints, whereas the incremental improvement gained by incorporating ECG and laboratory inputs beyond demographic information varied substantially across endpoints. Across the nine exploratory aggregated outcomes, Kaplan–Meier curves showed clear separation among model-score tertiles. Discrimination was highest for dementia (C-index, 0.891) and heart failure (C-index, 0.857). These findings support the feasibility of ECG-based longitudinal risk prediction across multiple diseases. External validation and competing-risk analyses are required to assess generalisability and clinical utility.

## Introduction

The routine 12-lead electrocardiogram (ECG) is inexpensive, non-invasive and embedded in clinical care. Beyond conventional assessment of rhythm, conduction and morphology, AI-enabled analysis can identify waveform features that are not apparent on visual interpretation. Deep-learning models detect cardiac abnormalities and subclinical ventricular dysfunction^1–3^, while cross-sectional studies have extended ECG-based detection to non-cardiac conditions^4–6^. Recent phenome-wide analyses have further linked learned ECG representations to prevalent and incident diseases across diverse organ systems^7^. Together, these studies broaden the information accessible from the ECG, but detecting existing disease or identifying population-level associations is distinct from estimating an individual’s future disease risk.

Using ECGs to predict future disease is a distinct and less studied objective. Longitudinal AI-ECG models have predicted mortality and broader cardiovascular risk, including atherosclerotic cardiovascular disease and major adverse cardiovascular events^8–10^. Condition-specific models have focused particularly on incident atrial fibrillation and heart failure^11–13^, and ECG-based models have also predicted ventricular arrhythmias and sudden cardiac death^14^. Most studies, however, have examined individual outcomes or small sets of predominantly cardiovascular endpoints. It therefore remains unclear whether a unified model can generate outcome-specific, time-to-event risk estimates for multiple future diseases across organ systems. Addressing this question requires a common cohort and survival framework that accommodates censored follow-up and distinguishes absolute model performance from the incremental prognostic information contributed by ECG waveforms and other clinical inputs.

Here we developed ECG-RISK, a multitask survival model integrating raw 12-lead ECGs with demographic and routinely collected laboratory data. Using linked MIMIC-IV^15^ data from 86,673 patients and a held-out test set, we modelled 67 incident ICD-10 endpoints, comprising 57 endpoints assigned to six focal organ systems and 10 additional endpoints retained for broader outcome coverage. We assessed endpoint-and organ-level discrimination, evaluated ECG waveforms as a standalone input, compared ECG-RISK with demographic and other input-matched models, and examined longitudinal risk stratification for nine representative aggregated clinical outcomes. By placing these endpoints within a common time-to-event framework, we sought to define both the breadth and heterogeneity of future disease information captured by routine ECGs, rather than to posit a universal ECG signature or a clinically deployable screening tool.

## Results

### Study cohort and characteristics

The study included 86,673 patients, of whom 73,672 formed the development cohort and 13,001 formed the held-out test set (**Fig. 1a**). Median age was 52 years (IQR, 36–65), and 49,235 patients (56.8%) were female. Demographic characteristics and laboratory-data availability and distributions were broadly similar between cohorts (**Table 1 and Supplementary Table 1**).

**Fig. 1.**
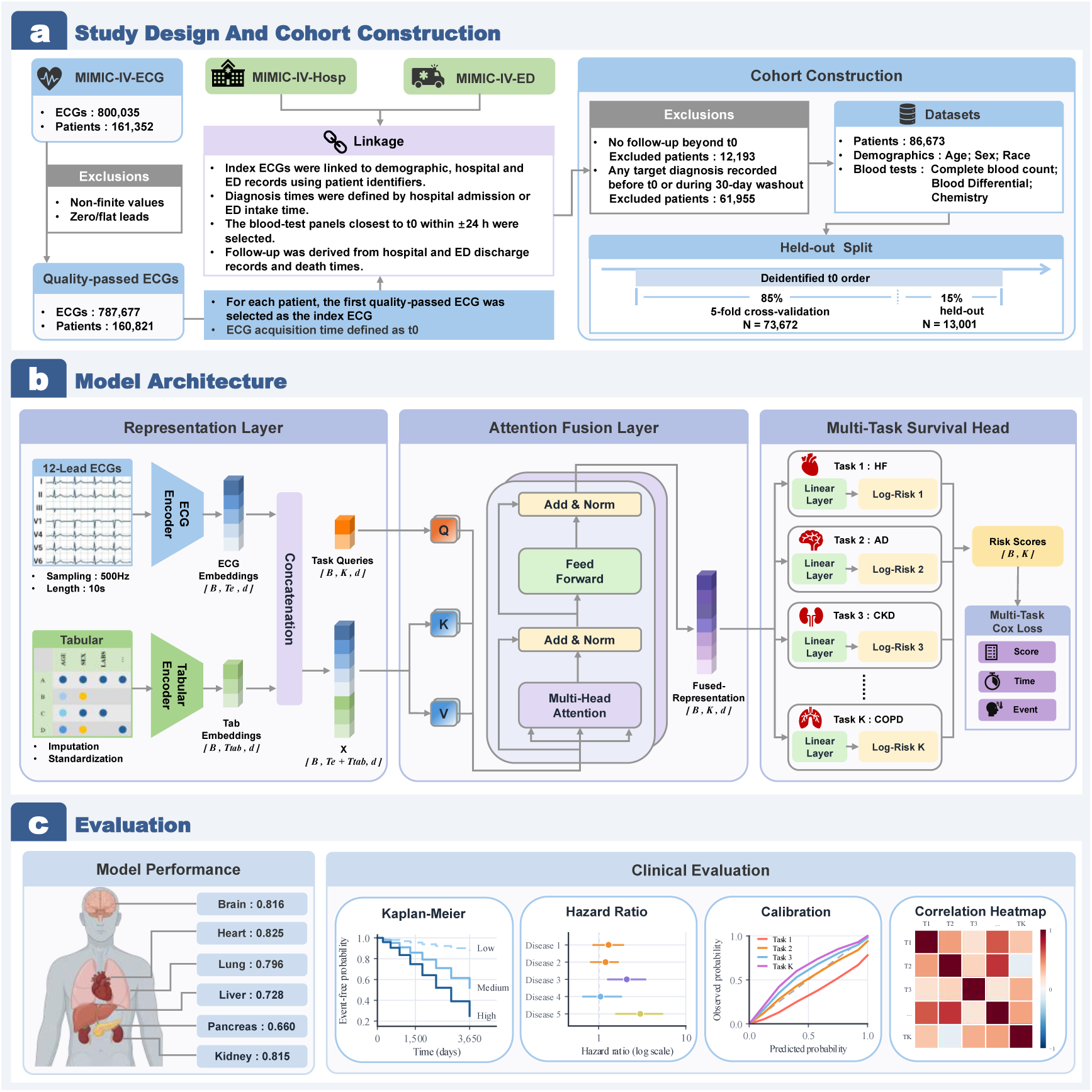
Overview of the study design and ECG-RISK framework. **a**, Linked MIMIC-IV resources were used to identify the index ECG, construct the incident disease cohort, define ICD-10 endpoints and assign patients to the development cohort or held-out test set. **b**, ECG waveforms and clinical variables were encoded separately, integrated through multi-head attention and passed to 67 endpoint-specific Cox survival heads. **c**, Evaluation included organ-and endpoint-level discrimination and, for nine representative aggregated clinical outcomes, Kaplan–Meier risk stratification, adjusted hazard ratios, calibration and risk-score correlation analyses.

**Table 1.** Cohort characteristics.

| Characteristics | Development | Test | Overall |
| --- | --- | --- | --- |
| **Patients, n** | 73,672 | 13,001 | 86,673 |
| **Age, median (IQR), years** | 52 (36-65) | 53 (38-65) | 52 (36-65) |
| **Age group, n (%)** |  |  |  |
| <40 years | 21,976 (29.8%) | 3,489 (26.8%) | 25,465 (29.4%) |
| 40-49 years | 11,058 (15.0%) | 2,141 (16.5%) | 13,199 (15.2%) |
| 50-59 years | 14,719 (20.0%) | 2,846 (21.9%) | 17,565 (20.3%) |
| 60-69 years | 12,828 (17.4%) | 2,198 (16.9%) | 15,026 (17.3%) |
| 70-79 years | 7,933 (10.8%) | 1,416 (10.9%) | 9,349 (10.8%) |
| 80-89 years | 4,222 (5.7%) | 739 (5.7%) | 4,961 (5.7%) |
| ≥90 years | 925 (1.3%) | 171 (1.3%) | 1,096 (1.3%) |
| Unknown | 11 (<0.1%) | 1 (<0.1%) | 12 (<0.1%) |
| **Sex, n (%)** |  |  |  |
| Female | 41,590 (56.5%) | 7,645 (58.8%) | 49,235 (56.8%) |
| Male | 32,071 (43.5%) | 5,355 (41.2%) | 37,426 (43.2%) |
| Unknown | 11 (<0.1%) | 1 (<0.1%) | 12 (<0.1%) |
| **Race, n (%)** |  |  |  |
| White | 45,552 (61.8%) | 7,837 (60.3%) | 53,389 (61.6%) |
| Black | 12,712 (17.3%) | 2,559 (19.7%) | 15,271 (17.6%) |
| Asian | 3,428 (4.7%) | 588 (4.5%) | 4,016 (4.6%) |
| Hispanic or Latino | 5,337 (7.2%) | 1,027 (7.9%) | 6,364 (7.3%) |
| Other | 6,643 (9.0%) | 990 (7.6%) | 7,633 (8.8%) |
| **Follow-up, n (%)** |  |  |  |
| ≥ 1 year | 38,308 (52.0%) | 7,546 (58.0%) | 45,854 (52.9%) |
| ≥ 3 years | 28,642 (38.9%) | 5,723 (44.0%) | 34,365 (39.6%) |
| ≥ 5 years | 20,440 (27.7%) | 4,053 (31.2%) | 24,493 (28.3%) |
| ≥ 10 years | 5,771 (7.8%) | 1,147 (8.8%) | 6,918 (8.0%) |
| **No events, n (%)** | **53,638 (72.8%)** | **9,180 (70.6%)** | **62,818 (72.5%)** |
| **At least 1 event, n (%)** | **20,034 (27.2%)** | **3,821 (29.4%)** | **23,855 (27.5%)** |
| **At least 2 events, n (%)** | **13,640 (18.5%)** | **2,609 (20.1%)** | **16,249 (18.7%)** |

Data are n (%) unless otherwise indicated; age is reported as median (IQR). Follow-up categories are cumulative.
Events refer to distinct incident ICD-10 endpoints recorded during follow-up. “Other” includes unknown or
unrecorded race or ethnicity. Percentages may not sum to 100% owing to rounding. IQR, interquartile range.

Follow-up extended for at least 1 year in 45,854 patients (52.9%), at least 5 years in 24,493 (28.3%) and at least 10 years in 6,918 (8.0%). Across the 67 individual ICD-10 endpoints, 23,855 patients (27.5%) experienced at least one incident endpoint and 16,249 (18.7%) experienced at least two. This combination of longitudinal observation and outcome accrual enabled evaluation of incident disease risk across organ systems, although rare endpoints and later time horizons were supported by fewer observations.

### ECG-RISK provides outcome-specific discrimination across organ systems

ECG-RISK provided outcome-specific discrimination across the six focal organ systems in the held-out test set (**Fig. 2 and Table 2**). Organ-level C-indices were highest for heart (0.825; 95% CI, 0.816 to 0.832), brain (0.816; 0.799 to 0.831) and kidney disease (0.815; 0.805 to 0.825), followed by lung disease (0.796; 0.780 to 0.812). Discrimination was lower for liver (0.728; 0.703 to 0.751) and pancreatic disease (0.660; 0.623 to 0.694). These findings showed that a common modelling framework could discriminate incident outcomes beyond the cardiovascular system, but performance differed substantially across disease domains.

**Fig. 2.**
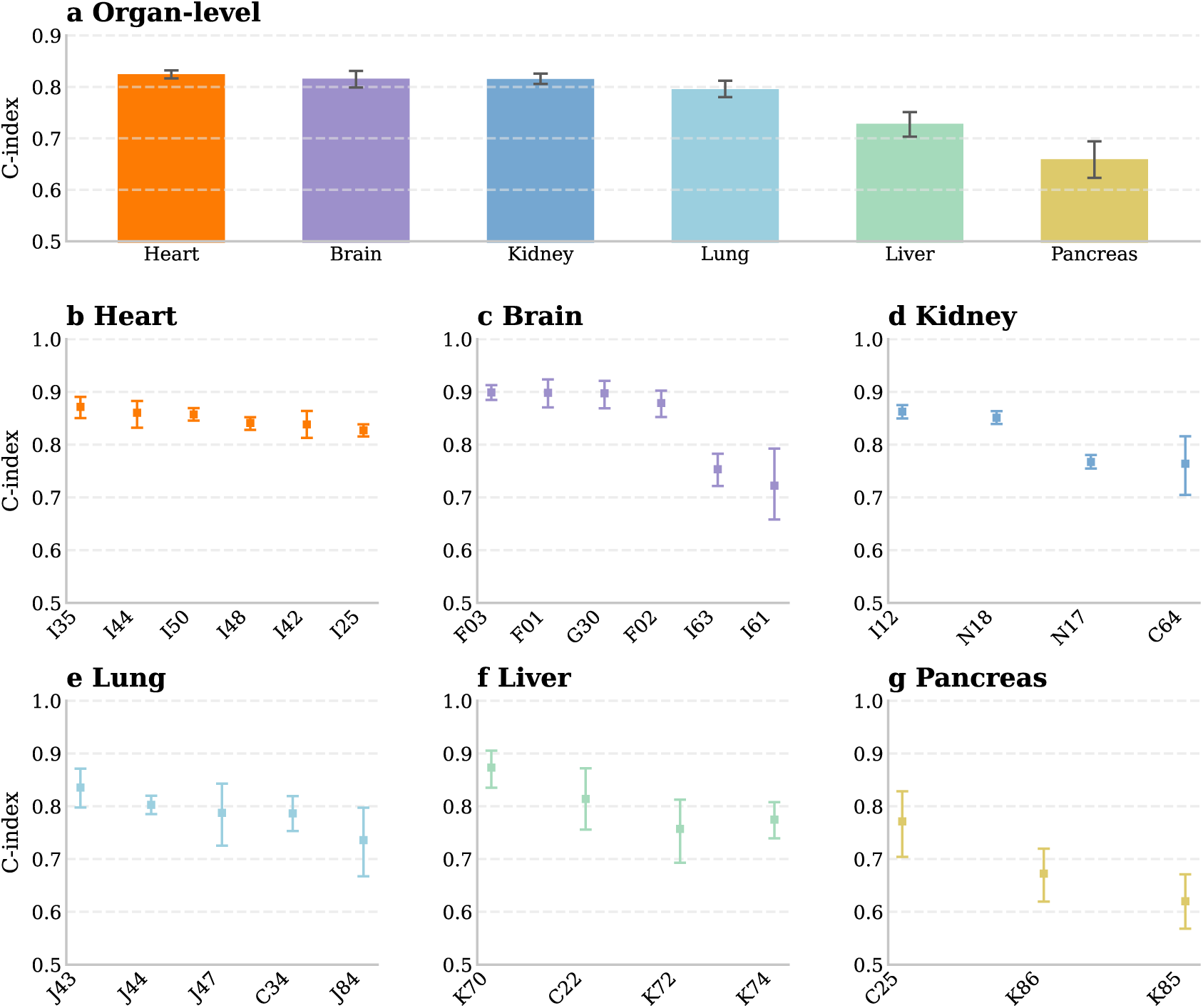
ECG-RISK discrimination across organ systems and incident disease endpoints. **a**, Organ-level C-indices for the six focal organ systems in the held-out test set, calculated as event-weighted averages of endpoint-level C-indices. **b–g**, C-indices for representative incident disease endpoints involving the heart (**b**), brain (**c**), kidney (**d**), lung (**e**), liver (**f**) and pancreas (**g**). Points and bars indicate C-index estimates, and error bars indicate 95% confidence intervals estimated using 1,000 patient-level bootstrap resamples. ICD-10 prefixes are shown on the horizontal axes. Complete results for all 67 endpoints are provided in **Supplementary Table 3**.

**Table 2.**
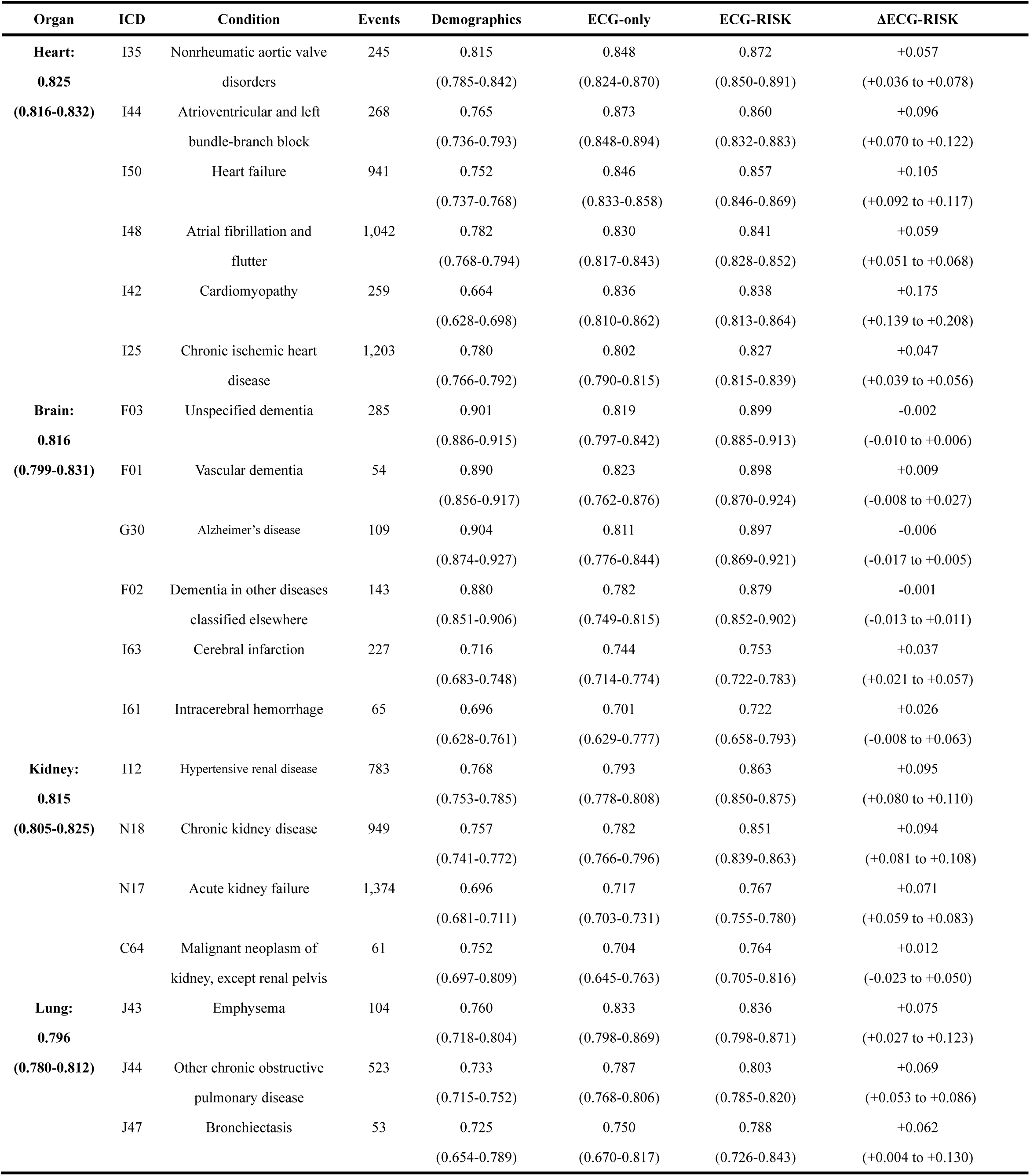
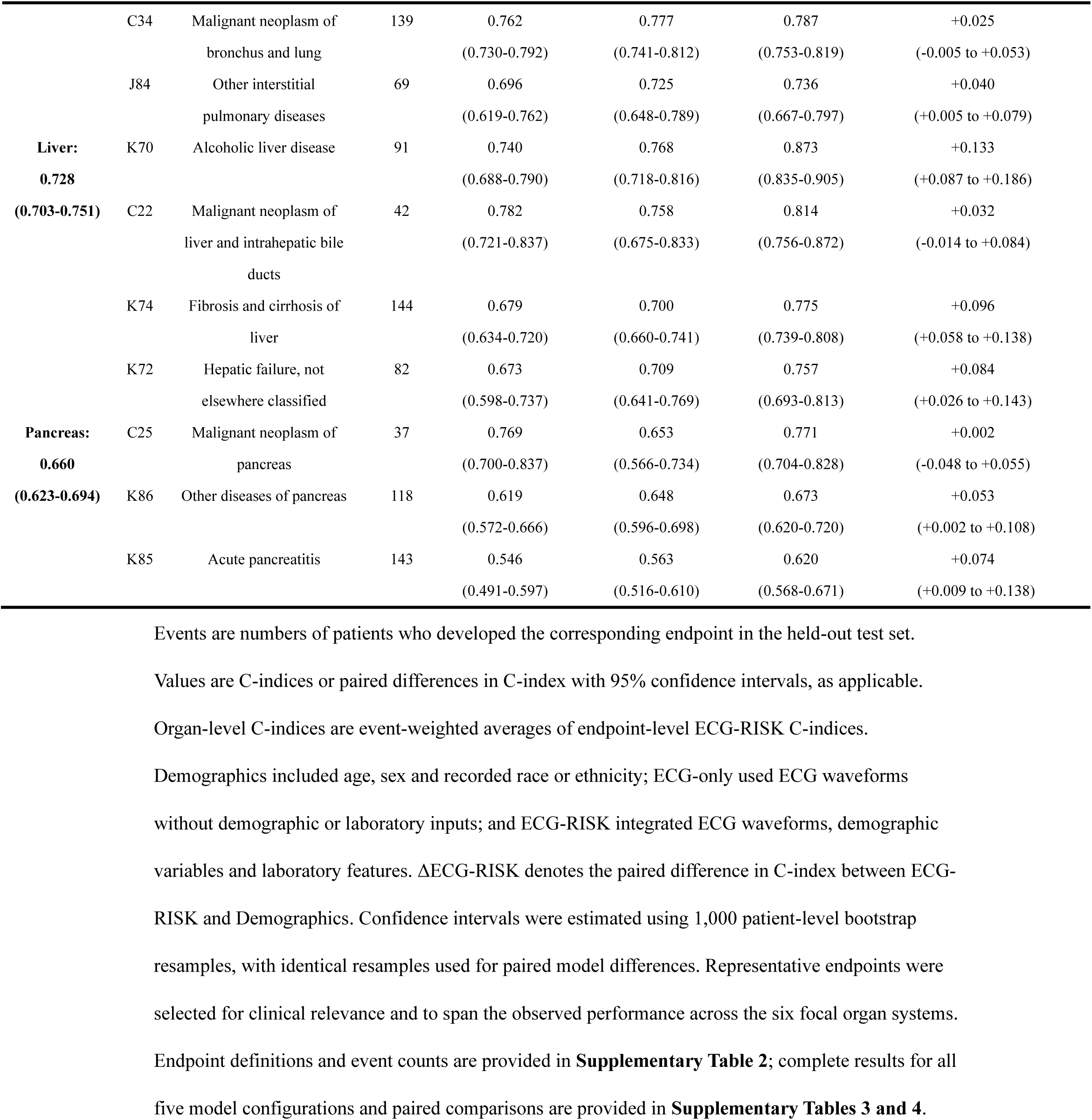
ECG-RISK discrimination at the organ and representative endpoint levels in the held-out test set.

High endpoint-level discrimination was not confined to cardiac disease. ECG-RISK achieved C-indices greater than 0.80 for all six displayed heart endpoints and for selected brain, kidney, lung and liver endpoints. These included unspecified dementia (0.899), vascular dementia (0.898), hypertensive renal disease (0.863), chronic kidney disease (0.851), emphysema (0.836) and alcoholic liver disease (0.873). Discrimination was more moderate for cerebrovascular and several other non-cardiac endpoints and was lowest for pancreatic diseases. Thus, high absolute performance extended across organ systems but remained concentrated in selected diseases.

To contextualize the performance of the full model, we evaluated an ECG-only comparator. ECG-only achieved high discrimination for atrioventricular and left bundle-branch block (C-index, 0.873), nonrheumatic aortic valve disorders (0.848), heart failure (0.846), cardiomyopathy (0.836) and emphysema (0.833), but showed less consistent performance across dementia, pancreatic and cancer endpoints. Relative to Demographics, ECG-RISK produced its largest paired improvements for cardiomyopathy (ΔC-index, +0.175; 95% CI, +0.139 to +0.208), alcoholic liver disease (+0.133; +0.087 to +0.186), heart failure (+0.105; +0.092 to +0.117) and hypertensive renal disease (+0.095; +0.080 to +0.110). In contrast, little or no improvement was observed for the displayed dementia and cancer endpoints. These comparisons showed that the value of the added ECG and laboratory inputs was endpoint specific rather than uniform across the disease spectrum. Results for the remaining comparator configurations and paired differences are provided in **Supplementary Tables 3 and 4.**

### Exploratory longitudinal risk stratification across aggregated outcomes

To examine whether endpoint-specific predictions yielded coherent longitudinal risk gradients, we performed exploratory analyses of nine aggregated clinical outcomes (**Table 3**). Discrimination was highest for dementia (C-index, 0.891; 95% CI, 0.877 to 0.904) and heart failure (0.857; 0.846 to 0.869), followed by ischemic heart disease (0.816; 0.804 to 0.826) and valvular heart disease (0.813; 0.793 to 0.833). More moderate discrimination was observed for kidney failure, chronic obstructive pulmonary disease, liver failure and stroke, whereas pancreatitis had the lowest C-index (0.627; 0.588 to 0.665). These analyses recapitulated the cross-domain heterogeneity observed at the individual-endpoint level.

**Table 3.** Discrimination of ECG-RISK for representative aggregated clinical outcomes in the held-out test set.

| Condition | ICD-10 prefix | Events, n | C-index (95% CI) |
| --- | --- | --- | --- |
| Heart failure | I50 | 941 | 0.857 (0.846-0.869) |
| Ischemic heart disease | I20, I21, I22, I23, I24, I25 | 1,334 | 0.816 (0.804-0.826) |
| Valvular heart disease | I34, I35, I36, I37, I38 | 535 | 0.813 (0.793-0.833) |
| Stroke | I60, I61, I62, I63 | 304 | 0.722 (0.693-0.750) |
| Dementia | F01, F02, F03, G30 | 392 | 0.891 (0.877-0.904) |
| Liver failure | K72 | 82 | 0.757 (0.693-0.813) |
| Kidney failure | N17, N18, N19 | 1,656 | 0.783 (0.771-0.795) |
| COPD | J41, J42, J43, J44 | 575 | 0.760 (0.739-0.779) |
| Pancreatitis | K85, K86 | 221 | 0.627 (0.588-0.665) |

Events are counts in the held-out test set. Aggregated clinical outcomes were defined by grouping clinically related ICD-10 prefixes. An event was defined as the first occurrence of any component endpoint, and the corresponding risk score was the maximum component endpoint-specific log-risk score. Values are C-indices (95% CIs). COPD, chronic obstructive pulmonary disease.

Outcome-specific ECG-RISK scores separated patients into ordered risk groups (**Fig. 3**). Across all nine aggregated outcomes, patients in the highest score tertile had lower incident-free probabilities than those in the intermediate and lowest tertiles. Separation was most pronounced for heart failure, ischemic heart disease and kidney failure, and was also evident for dementia, valvular heart disease and chronic obstructive pulmonary disease. More modest separation was observed for stroke, liver failure and pancreatitis. These descriptive tertile analyses showed graded risk stratification across all nine outcomes, although the magnitude of separation varied.

**Fig. 3.**
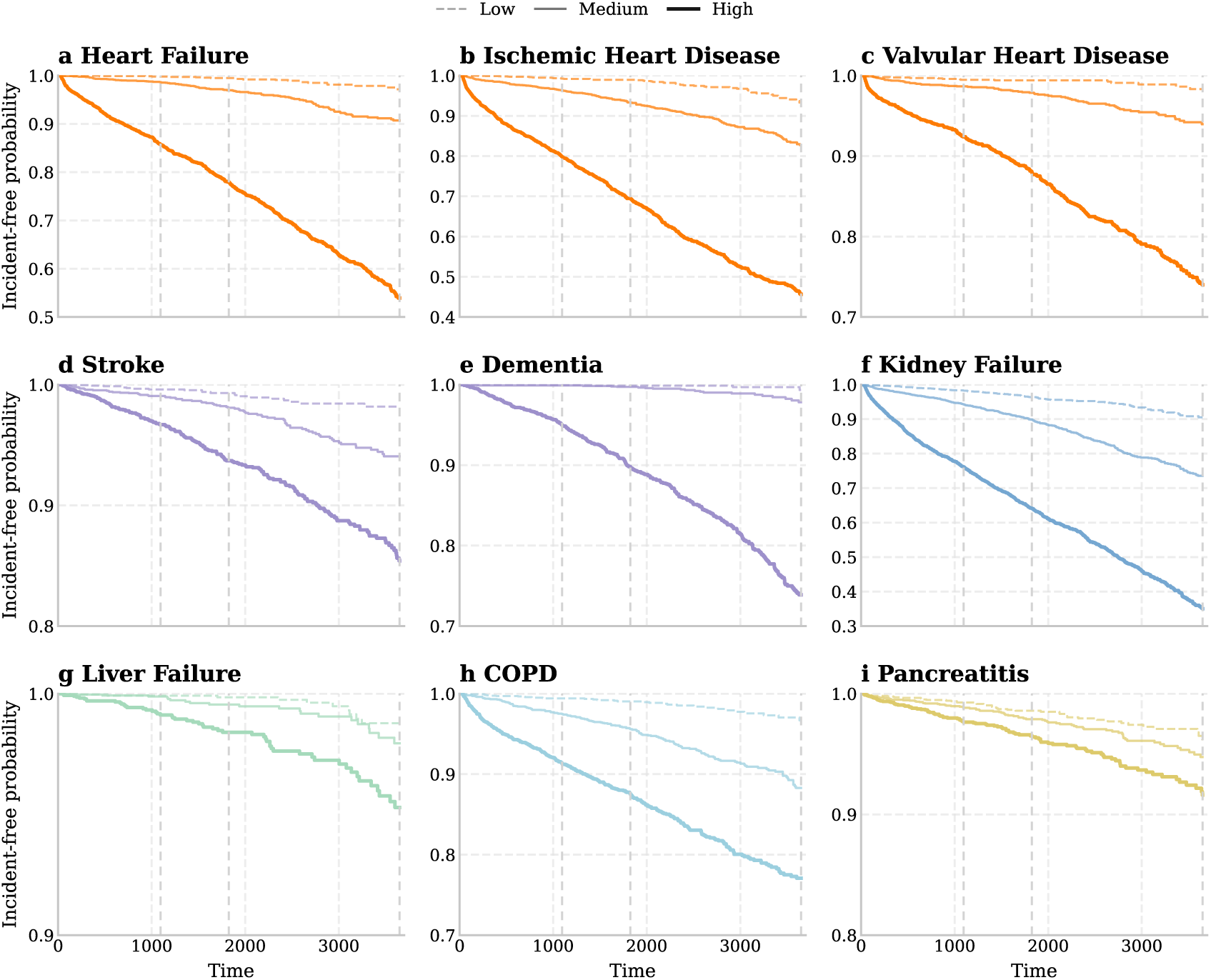
Risk stratification by ECG-RISK across representative aggregated clinical outcomes. Kaplan–Meier estimates of incident-free probability, S(t), are shown for nine representative aggregated clinical outcomes in the held-out test set. Patients were stratified separately for each outcome into tertiles of the corresponding ECG-RISK log-risk score, with line styles denoting low-, medium-and high-risk groups. Vertical grey dashed lines indicate the 3-, 5-and 10-year horizons. Time was measured from the index ECG.

Continuous score analyses were consistent with these graded associations (**Fig. 4**). After adjustment for age and sex, hazard ratios per 1-s.d. increase in the outcome-specific ECG-RISK score ranged from 1.48 for pancreatitis (95% CI, 1.28 to 1.71) to 4.36 for heart failure (3.99 to 4.77), with all confidence intervals excluding 1. Within the cause-specific censoring framework, agreement between predicted and observed event probabilities was generally closer at 3 and 5 years than at 10 years, with greater deviations at higher predicted probabilities and later horizons (**Supplementary Fig. 1**). Exploratory score correlations showed shared variation among model outputs but were not interpreted as evidence of disease co-occurrence (**Supplementary Figs. 2 and 3**). Together, these analyses showed that ECG-RISK scores consistently ordered patients by subsequent risk across cardiac and non-cardiac outcomes, although the strength and calibration of this stratification varied across outcomes.

**Fig. 4.**
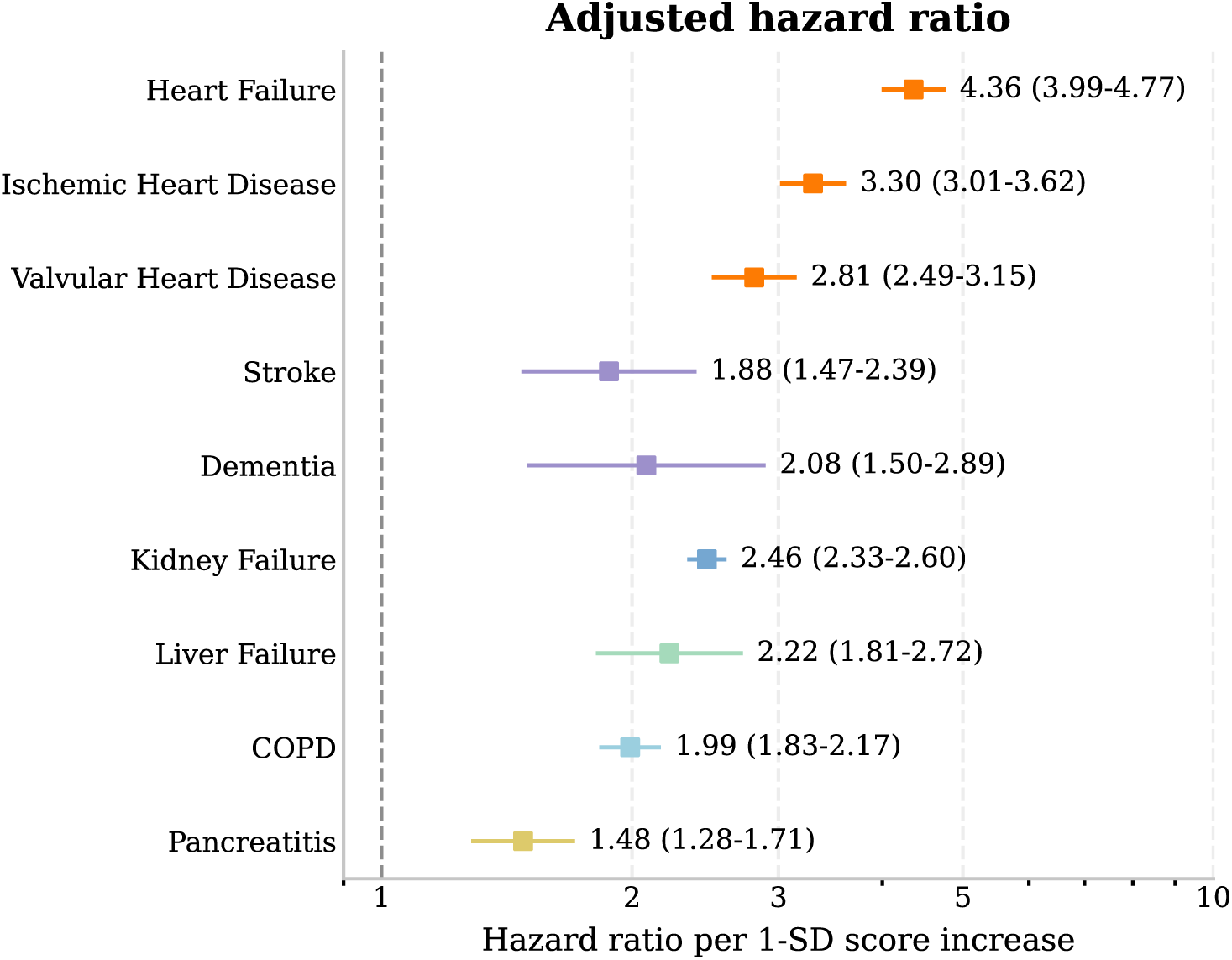
Adjusted hazard ratios for ECG-RISK scores across representative aggregated clinical outcomes. Squares and horizontal lines indicate hazard ratios and 95% confidence intervals, respectively, from separate Cox proportional hazards models adjusted for age and sex in the held-out test set. Estimates are reported per 1-s.d. increase in the outcome-specific ECG-RISK log-risk score. The vertical dashed line indicates a hazard ratio of 1.

## Discussion

This study extends AI-enabled ECG analysis from contemporaneous disease detection and disease-specific forecasting towards unified longitudinal risk estimation across organ systems. ECG-RISK modelled 67 incident ICD-10 endpoints within a common time-to-event framework using a single index ECG and baseline clinical data, while ECG-only quantified prognostic information from the waveform alone. In the single-centre held-out cohort, discrimination extended beyond cardiovascular outcomes but varied substantially among diseases, as did improvement over demographic risk. The central finding is therefore not a universal ECG signature of future disease, but the feasibility of generating outcome-specific risk estimates for multiple incident diseases within a common modelling framework.

Prior longitudinal AI-ECG studies have focused on mortality^9,16^ or individual cardiovascular outcomes^11–13^, whereas non-cardiac applications have largely centred on cross-sectional diagnosis^4,6^. A recent phenome-wide analysis showed that learned ECG representations were associated with hundreds of incident diseases^7^, providing evidence that the ECG carries broad prognostic information. ECG-RISK addresses a complementary but distinct question: whether an outcome-supervised multitask survival model can generate discriminative, endpoint-specific scores across multiple future diseases within a common longitudinal design. The contrast between high absolute discrimination for dementia and little improvement of ECG-RISK over Demographics illustrates that final-model performance does not necessarily imply substantial information from the added ECG and laboratory inputs. Conversely, gains were larger for selected cardiac, respiratory, cerebrovascular, kidney and liver endpoints, and the ECG-only comparator retained substantial discrimination for selected cardiac and respiratory diseases. This heterogeneity argues for endpoint-specific evaluation against appropriate baselines rather than treating AI-ECG as a generic disease-prediction tool.

Cross-organ disease risk can share predictive structure across physiological systems and longitudinal health records^17,18^. Autonomic regulation, conduction, hemodynamic stress, cardiopulmonary interactions and metabolic or electrolyte disturbances can influence ECG morphology and may be associated with subsequent clinical trajectories^19,20^. However, model scores may also encode age, comorbidity, healthcare use and clinical measurement patterns through the model inputs and training data^21^. Correlations among endpoint scores should therefore be interpreted as shared predictive structure, not disease co-occurrence or causal relationships. ECG-RISK is best viewed as an index-encounter prognostic model for future EHR-recorded morbidity, rather than a mechanistic biomarker or a validated screening test.

Several limitations constrain the interpretation of these findings. First, the held-out test set was drawn from the same institution and data infrastructure, and subject-specific date shifting precluded true calendar-time temporal validation; neither temporal generalizability nor external transportability was established. Second, outcomes were derived from hospital and emergency department diagnosis codes, such that event times represented first recorded diagnoses rather than verified biological onset. EHR-observed follow-up may also have missed outpatient diagnoses and events occurring outside the data system. Third, the 30-day washout reduced contamination from prevalent or index-encounter disease but could not exclude undocumented prior disease. The common exclusion of any prior diagnosis among the 67 target endpoints also produced a selected disease-free cohort and may limit generalizability. Rare endpoints and the smaller number of observations at later horizons reduced the precision of some estimates, particularly for long-term calibration. Performance across demographic and clinical subgroups was not systematically evaluated.

Additional methodological constraints warrant emphasis. Death was treated as censoring rather than as a competing event, so the reported event probabilities do not estimate real-world cumulative incidence when mortality is informative. The primary ECG-RISK versus Demographics comparison jointly added ECG and laboratory features and therefore did not isolate the incremental contribution of ECG beyond complete clinical data. Laboratory measurements obtained within 24 h before or after the index ECG further define ECG-RISK as an index-encounter model combining ECG and clinical data rather than an ECG-only screening model. Finally, the aggregated clinical outcomes were exploratory, were derived as the maximum of component endpoint scores and were not independently trained; because this maximum is scale dependent, these analyses should be interpreted as descriptive risk stratification.

The widespread availability and low marginal cost of routine ECG acquisition make it attractive for opportunistic risk assessment. By demonstrating ordered risk across cardiac and non-cardiac outcomes, this study provides a proof of concept for ECG-informed, multi-disease longitudinal risk estimation; it does not establish clinical utility. Before clinical evaluation or implementation, ECG-RISK requires multicentre external validation, population-specific recalibration, subgroup and fairness evaluation, and comparison with established disease-specific risk tools^21^. Subsequent live clinical evaluations should determine whether endpoint-specific risk estimates alter clinical decisions or patient outcomes and should report safety, workflow and human-factor effects^22^. Within these boundaries, our findings motivate further evaluation of routine ECGs as one component of longitudinal risk assessment across multiple organ systems.

## Methods

### Data sources and cohort construction

We conducted a retrospective cohort study using the de-identified MIMIC-IV v3.1^15^, MIMIC-IV-ED v2.2^23^ and MIMIC-IV-ECG v1.0^24^ databases. The linked data included 12-lead ECG waveforms, hospital and emergency department encounters, diagnosis codes, demographic variables and laboratory measurements. The study was conducted under the MIMIC data-use agreement. The underlying data collection and sharing procedures were approved by the Beth Israel Deaconess Medical Center Institutional Review Board with a waiver of informed consent. MIMIC-IV applies a single subject-specific date shift, preserving within-patient intervals but not calendar-time comparability between patients.

The ECG database contained 800,035 recordings from 161,352 patients. Recordings with missing required leads, unreadable or non-finite values, zero-valued waveforms or near-flat leads were excluded. Leads were reordered into the standard 12-lead sequence, and the first quality-passed ECG for each patient was selected as the index ECG. Its acquisition time was defined as *t*_0_. This yielded 787,677 quality-passed ECGs from 160,821 patients before outcome-based cohort construction.

Patients without EHR-observed follow-up beyond *t*_0_ were excluded. We also excluded patients with any target diagnosis recorded before *t*_0_ or during the 30-day washout period from *t*_0_ through day 30. The final analytic cohort comprised 86,673 patients. This yielded a common cohort without recorded prior diagnoses for any of the 67 target endpoints.

### Endpoints and follow-up

Incident outcomes were defined using three-character ICD-10 prefixes. ICD-9 diagnoses were mapped to ICD-10 before prefix extraction. Hospital diagnoses were assigned the corresponding admission time, whereas emergency department diagnoses were assigned the encounter intake time.

We modelled 67 individual ICD-10 endpoints, including 57 endpoints assigned to six focal organ systems and 10 additional endpoints retained for broader outcome coverage. Detailed endpoint definitions, organ-system assignments and event counts are provided in **Supplementary Table 2**. For each endpoint, an incident event was defined as the first corresponding diagnosis recorded after the 30-day washout and before censoring.

For each endpoint, time-to-event was measured from *t*_0_ to the first qualifying diagnosis. Patients without an event were censored at the earliest of death, 10 years after *t*_0_, or the last observed EHR contact, defined as the later of the final hospital and emergency department discharge times. Death was treated as a censoring event in these cause-specific analyses.

### Model inputs and architecture

Eligible ECGs were 10-s, 12-lead recordings sampled at 500 Hz. Each lead was independently centred and scaled to unit variance. Clinical inputs comprised age, sex and recorded race or ethnicity. Laboratory inputs were obtained from complete blood count, blood differential and chemistry panels. For each panel, the row closest in absolute time to *t*_0_ within 24 h before or after the index ECG was selected. Candidate laboratory variables with more than 90% missingness were excluded before model fitting. Retained laboratory variables were accompanied by missingness indicators and the signed time interval between *t*_0_ and the selected panel. Continuous tabular variables were median-imputed and standardised using parameters estimated from the training patients in each fold. Categorical variables were one-hot encoded, with separate indicators for missing values. The fitted transformations were applied without refitting to the corresponding validation fold and test set. In accordance with MIMIC-IV de-identification, patients older than 89 years were represented in a single grouped age category.

ECG-RISK is a multimodal multitask survival network comprising an ECG encoder, a tabular projection module, endpoint-specific task queries and independent survival heads (**Fig. 1**). ECG waveforms were encoded using a previously described convolutional–Transformer architecture^25^. The encoder was initialized without pretrained weights and produced a sequence of 128-dimensional ECG embeddings. Tabular variables were projected into eight 128-dimensional tabular embeddings. The ECG and tabular embedding sequences were concatenated along the sequence dimension to form a shared multimodal context. Learned endpoint-specific task queries interacted with this context through two cross-attention blocks, and the resulting representations were passed to 67 independent linear survival heads to generate endpoint-specific log-risk scores.

### Model development and comparator models

Patients were ordered by *t*_0_. The final 15% in this deidentified ordering were reserved as the held-out test set (*n* = 13,001), and the remaining 85% formed the development cohort (*n* = 73,672). The development cohort was randomly divided into five folds. For each fold, four folds were used for training and one for validation and out-of-fold prediction. Fold-specific models were applied to the held-out test set without updating model parameters or selecting checkpoints using test-set data. Test-set log-risk scores were averaged across the five fold-specific models. Because MIMIC-IV shifts dates independently for each patient, this ordering does not represent calendar-time temporal validation; it was used only to define a deterministic patient-level holdout.

The multitask objective used a mini-batch approximation to the negative Cox partial log-likelihood^26^, with endpoint losses averaged across tasks containing an observed event in the corresponding batch. Models were trained for 15 epochs using AdamW with a learning rate of 1 × 10^−4^, weight decay of 1 × 10^−4^, a batch size of 64 and gradient-norm clipping at 5.0. During the first two epochs, the ECG encoder was frozen before joint optimization of all model components. Checkpoints were selected using the unweighted mean validation C-index across endpoints with at least 50 events in the corresponding training partition.

Five model configurations were trained using identical cohort splits and optimization procedures. Demographics included age, sex and recorded race or ethnicity; ECG-only used ECG waveforms alone; ECG-demographics combined ECG waveforms with demographic variables; Demographics-laboratory combined demographic variables with laboratory features, timing variables and missingness indicators; and ECG-RISK included ECG waveforms, demographic variables and laboratory features. The Demographics and Demographics-laboratory models used a tabular multitask network comprising LayerNorm, two linear–GELU–dropout blocks and 67 endpoint-specific outputs. ECG-only used the same ECG encoder, task-query attention blocks and endpoint-specific survival heads as ECG-RISK but omitted all tabular embeddings. ECG-demographics used the same multimodal task-query architecture as ECG-RISK but omitted laboratory inputs.

Discrimination was assessed using Harrell’s C-index^27^ in the held-out test set. Ninety-five percent confidence intervals were estimated from 1,000 patient-level bootstrap resamples.

The primary paired difference reported in **Table 2** was defined as Δ_ECG−RISK_ = *C*_ECG−RISK_ − *C*_Demographics_. Secondary paired comparisons reported in **Supplementary Table 4** included Δ_Labora*t*ory_ = *C*_Demographics−labora*t*ory_ − *C*_Demographics_ and Δ_ECG_ = *C*_ECG−d*emographics*_ − *C*_Demographics_. Identical patient-level bootstrap samples were used for the two models in each comparison. Organ-level C-indices were calculated as event-weighted averages of endpoint-level C-indices and were treated as descriptive summaries rather than estimates for pooled organ-level outcomes.

Representative endpoints in **Table 2** were selected to cover clinically relevant conditions across the six focal organ systems and the range of observed performance. Complete results for all 67 endpoints, including those with few or no events in the held-out test set, are reported in the Supplementary Information. Analyses across endpoints were estimation-focused, and no adjustment for multiple comparisons was applied.

### Aggregated outcomes and supplementary analyses

Nine clinically representative aggregated outcomes were evaluated: heart failure, ischemic heart disease, valvular heart disease, stroke, dementia, liver failure, kidney failure, chronic obstructive pulmonary disease and pancreatitis. Each outcome grouped related ICD-10 prefixes, and an aggregated event was defined as the first occurrence of any component endpoint. No separate model was trained for these outcomes. Within each fold, the aggregated score was defined as the maximum of the component endpoint-specific scores and then averaged across fold-specific models. These aggregated analyses were exploratory. Because the maximum of component log-risk scores is scale dependent, the results should be interpreted as descriptive risk stratification rather than as validation of independently trained composite outcomes.

Patients were stratified into tertiles of each aggregated score for Kaplan–Meier analyses. Hazard ratios were estimated per 1-s.d. increase in the standardised aggregated score using Cox proportional hazards models adjusted for age and sex. Calibration under the cause-specific censoring framework was evaluated at 3, 5 and 10 years using fold-specific penalized Cox models fitted to standardised out-of-fold development scores and applied without refitting to the corresponding test-set scores. Observed net event probabilities were estimated as 1 − *S*(*t*) using the Kaplan–Meier estimator. Spearman correlations among representative endpoint-level and aggregated risk scores were reported as descriptive analyses. Calibration results and score-correlation matrices are provided in **Supplementary Fig. 1** and **Supplementary Figs. 2 and 3**, respectively. Because death was censored, these quantities do not represent competing-risk cumulative incidence.

## Data Availability

This study used the publicly available, de-identified MIMIC-IV v3.1, MIMIC-IV-ED v2.2 and MIMIC-IV-ECG v1.0 databases, which are hosted on PhysioNet. Access to these datasets is controlled by PhysioNet and requires completion of the required training, credentialing and data use agreement. The authors are not permitted to redistribute the raw patient-level data. The endpoint definitions and aggregate results needed to interpret the findings are provided in the Article and Supplementary Information.

## Author contributions

Y.Y. designed the methodology, developed the software, curated the data, performed the statistical analyses, prepared the figures and tables, and drafted the manuscript. Z.Z. contributed to data curation and visualization. X.T., Z.Y. and J.W. contributed to data curation, study design discussions and interpretation of the findings. Y.Z. conceived and supervised the study and reviewed and edited the manuscript. All authors reviewed and approved the final manuscript and agreed to be accountable for their contributions and the integrity of the work.

## Data Availability

The de-identified MIMIC-IV v3.1, MIMIC-IV-ED v2.2 and MIMIC-IV-ECG v1.0 databases are available through PhysioNet under a credentialed-access process requiring completion of the relevant training, credentialing and data-use agreement. The authors are not permitted to redistribute the raw patient-level data. Endpoint definitions and the summary data supporting the reported results are provided in the manuscript and Supplementary Information.

https://physionet.org/content/mimiciv/3.1/

https://physionet.org/content/mimic-iv-ed/2.2/

https://physionet.org/content/mimic-iv-ecg/1.0/

