## Supplementary Information for "ECG-based longitudinal risk prediction across diseases and organ systems"

### Supplementary Table 1 | Availability and distribution of laboratory variables in the development cohort and held-out test set.

| Laboratory panel | Laboratory variable | Unit | Development (n=73,672) available, n (%) | Development median (IQR) | Held-out test (n=13,001) available, n (%) | Held-out test median (IQR) |
|---|---|---|---|---|---|---|
| **Complete blood count** | Hematocrit | % | 49,437 (67.1%) | 39.4 (36.4-42.3) | 8,894 (68.4%) | 39.3 (36.3-42.2) |
| | Hemoglobin | g/dL | 49,121 (66.7%) | 13.3 (12.2-14.4) | 8,843 (68.0%) | 13.3 (12.2-14.3) |
| | Mean corpuscular hemoglobin | pg | 49,072 (66.6%) | 30.1 (28.7-31.3) | 8,835 (68.0%) | 30.0 (28.7-31.3) |
| | Mean corpuscular hemoglobin concentration | g/dL | 49,077 (66.6%) | 33.7 (32.8-34.6) | 8,835 (68.0%) | 33.7 (32.7-34.5) |
| | Mean corpuscular volume | fL | 49,074 (66.6%) | 89.0 (85.0-92.0) | 8,835 (68.0%) | 89.0 (85.0-92.0) |
| | Platelet count | 10^3/uL | 49,095 (66.6%) | 248 (204-299) | 8,842 (68.0%) | 249 (204-299) |
| | Red blood cell count | 10^6/uL | 49,076 (66.6%) | 4.45 (4.08-4.81) | 8,835 (68.0%) | 4.43 (4.09-4.81) |
| | Red cell distribution width | % | 49,067 (66.6%) | 13.2 (12.7-14.0) | 8,834 (67.9%) | 13.3 (12.7-14.0) |
| | White blood cell count | 10^3/uL | 49,107 (66.7%) | 7.8 (6.1-10.1) | 8,842 (68.0%) | 7.7 (6.1-9.9) |
| **Blood differential** | White blood cell count | 10^3/uL | 49,365 (67.0%) | 7.8 (6.1-10.1) | 8,874 (68.3%) | 7.7 (6.1-9.9) |
| | Neutrophils | % | 39,336 (53.4%) | 66.2 (57.1-76.1) | 7,114 (54.7%) | 65.7 (56.5-75.5) |
| | Lymphocytes | % | 39,346 (53.4%) | 24.8 (16.2-33.0) | 7,120 (54.8%) | 25.3 (16.7-33.7) |
| | Monocytes | % | 39,336 (53.4%) | 5.4 (4.1-7.0) | 7,114 (54.7%) | 5.4 (4.1-7.1) |
| | Eosinophils | % | 39,336 (53.4%) | 1.4 (0.6-2.5) | 7,114 (54.7%) | 1.5 (0.7-2.6) |
| | Basophils | % | 39,336 (53.4%) | 0.5 (0.3-0.7) | 7,114 (54.7%) | 0.5 (0.3-0.8) |
| | Absolute neutrophil count | 10^3/uL | 39,351 (53.4%) | 5.09 (3.63-7.29) | 7,117 (54.7%) | 5.00 (3.61-7.10) |

| | | | | | | |
|---|---|---|---|---|---|---|
| | Absolute lymphocyte count | 10^3/uL | 39,346 (53.4%) | 1.83 (1.32-2.42) | 7,120 (54.8%) | 1.85 (1.34-2.44) |
| | Absolute monocyte count | 10^3/uL | 39,336 (53.4%) | 0.42 (0.31-0.58) | 7,114 (54.7%) | 0.42 (0.31-0.58) |
| | Absolute eosinophil count | 10^3/uL | 39,336 (53.4%) | 0.11 (0.05-0.19) | 7,114 (54.7%) | 0.11 (0.06-0.20) |
| | Absolute basophil count | 10^3/uL | 39,336 (53.4%) | 0.04 (0.03-0.06) | 7,114 (54.7%) | 0.04 (0.03-0.06) |
| | Immature granulocytes | % | 12,032 (16.3%) | 0.4 (0.3-0.5) | 2,092 (16.1%) | 0.4 (0.3-0.5) |
| **Chemistry** | Albumin | g/dL | 9,788 (13.3%) | 4.3 (4.0-4.6) | 1,771 (13.6%) | 4.3 (4.0-4.6) |
| | Anion gap | mEq/L | 43,832 (59.5%) | 15.0 (13.0-17.0) | 7,861 (60.5%) | 15.0 (13.0-16.0) |
| | Bicarbonate | mEq/L | 43,873 (59.6%) | 25.0 (23.0-27.0) | 7,872 (60.5%) | 25.0 (23.0-27.0) |
| | Blood urea nitrogen | mg/dL | 45,620 (61.9%) | 14.0 (11.0-18.0) | 8,146 (62.7%) | 14.0 (11.0-18.0) |
| | Calcium | mg/dL | 14,252 (19.3%) | 9.2 (8.8-9.6) | 2,534 (19.5%) | 9.3 (8.9-9.6) |
| | Chloride | mEq/L | 44,330 (60.2%) | 103.0 (100.0-105.0) | 7,954 (61.2%) | 103.0 (100.0-105.0) |
| | Creatinine | mg/dL | 45,895 (62.3%) | 0.80 (0.70-1.00) | 8,191 (63.0%) | 0.80 (0.70-1.00) |
| | Glucose | mg/dL | 42,962 (58.3%) | 103.0 (92.0-122.0) | 7,743 (59.6%) | 102.0 (92.0-122.0) |
| | Sodium | mEq/L | 44,371 (60.2%) | 139.0 (137.0-141.0) | 7,960 (61.2%) | 139.0 (137.0-141.0) |
| | Potassium | mEq/L | 44,467 (60.4%) | 4.0 (3.8-4.3) | 7,972 (61.3%) | 4.0 (3.8-4.3) |

Laboratory availability was defined by an observed measurement within 24 h before or after the index ECG. For each laboratory panel, the closest eligible row to the index ECG was selected. Values are n (%) and median (IQR) among patients with observed measurements and were summarized before imputation and standardization. Only variables retained after the feature-availability screen are shown. IQR, interquartile range.

**Supplementary Table 2 | Definitions and event counts for the 67 individual ICD-10 endpoints.**

| Organ system | ICD-10 prefix | Condition | Development events, n | Held-out test events, n | Overall events, n |
|---|---|---|---|---|---|
| **Heart** | I05 | Rheumatic mitral valve diseases | 108 | 23 | 131 |
| | I06 | Rheumatic aortic valve diseases | 18 | 1 | 19 |
| | I07 | Rheumatic tricuspid valve diseases | 416 | 100 | 516 |
| | I08 | Multiple valve diseases | 875 | 165 | 1,040 |
| | I09 | Other rheumatic heart diseases | 74 | 14 | 88 |
| | I11 | Hypertensive heart disease | 1,769 | 354 | 2,123 |
| | I20 | Angina pectoris | 891 | 199 | 1,090 |
| | I21 | Acute myocardial infarction | 1,653 | 327 | 1,980 |
| | I22 | Subsequent myocardial infarction | 18 | 4 | 22 |
| | I23 | Certain current complications following acute myocardial infarction | 18 | 0 | 18 |
| | I24 | Other acute ischemic heart diseases | 474 | 118 | 592 |
| | I25 | Chronic ischemic heart disease | 6,629 | 1,203 | 7,832 |
| | I34 | Nonrheumatic mitral valve disorders | 1,407 | 300 | 1,707 |
| | I35 | Nonrheumatic aortic valve disorders | 1,477 | 245 | 1,722 |
| | I36 | Nonrheumatic tricuspid valve disorders | 117 | 26 | 143 |
| | I37 | Nonrheumatic pulmonary valve disorders | 24 | 5 | 29 |
| | I38 | Endocarditis, valve unspecified | 48 | 14 | 62 |
| | I42 | Cardiomyopathy | 1,282 | 259 | 1,541 |
| | I44 | Atrioventricular and left bundle-branch block | 1,380 | 268 | 1,648 |
| | I45 | Other conduction disorders | 915 | 180 | 1,095 |
| | I47 | Paroxysmal tachycardia | 1,497 | 319 | 1,816 |

| | | | | | |
|---|---|---|---|---|---|
| | I48 | Atrial fibrillation and flutter | 5,590 | 1,042 | 6,632 |
| | I50 | Heart failure | 4,907 | 941 | 5,848 |
| | I70 | Atherosclerosis | 955 | 178 | 1,133 |
| | I71 | Aortic aneurysm and dissection | 630 | 112 | 742 |
| | I73 | Other peripheral vascular diseases | 1,267 | 267 | 1,534 |
| **Brain** | C71 | Malignant neoplasm of brain | 83 | 14 | 97 |
| | F01 | Vascular dementia | 253 | 54 | 307 |
| | F02 | Dementia in other diseases classified elsewhere | 711 | 143 | 854 |
| | F03 | Unspecified dementia | 1,173 | 285 | 1,458 |
| | G20 | Parkinson's disease | 417 | 74 | 491 |
| | G30 | Alzheimer's disease | 500 | 109 | 609 |
| | G81 | Hemiplegia and hemiparesis | 573 | 112 | 685 |
| | I60 | Nontraumatic subarachnoid hemorrhage | 118 | 23 | 141 |
| | I61 | Intracerebral haemorrhage | 347 | 65 | 412 |
| | I62 | Other and unspecified nontraumatic intracranial hemorrhage | 240 | 38 | 278 |
| | I63 | Cerebral infarction | 1,153 | 227 | 1,380 |
| **Kidney** | C64 | Malignant neoplasm of kidney, except renal pelvis | 243 | 61 | 304 |
| | I12 | Hypertensive renal disease | 3,695 | 783 | 4,478 |
| | N17 | Acute kidney failure | 6,785 | 1,374 | 8,159 |
| | N18 | Chronic kidney disease (CKD) | 4,687 | 949 | 5,636 |
| | N19 | Unspecified kidney failure | 63 | 11 | 74 |
| **Lung** | C34 | Malignant neoplasm of bronchus and lung | 635 | 139 | 774 |
| | J41 | Simple and mucopurulent chronic bronchitis | 8 | 3 | 11 |
| | J42 | Unspecified chronic bronchitis | 45 | 9 | 54 |
| | J43 | Emphysema | 465 | 104 | 569 |

| | | | | | |
|---|---|---|---|---|---|
| | J44 | Other chronic obstructive pulmonary disease | 2,850 | 523 | 3,373 |
| | J47 | Bronchiectasis | 223 | 53 | 276 |
| | J84 | Other interstitial pulmonary diseases | 432 | 69 | 501 |
| **Liver** | C22 | Malignant neoplasm of liver and intrahepatic bile ducts | 283 | 42 | 325 |
| | K70 | Alcoholic liver disease | 509 | 91 | 600 |
| | K72 | Hepatic failure, not elsewhere classified | 472 | 82 | 554 |
| | K74 | Fibrosis and cirrhosis of liver | 833 | 144 | 977 |
| | K76 | Other diseases of liver | 1,929 | 394 | 2,323 |
| **Pancreas** | C25 | Malignant neoplasm of pancreas | 236 | 37 | 273 |
| | K85 | Acute pancreatitis | 635 | 143 | 778 |
| | K86 | Other diseases of pancreas | 618 | 118 | 736 |
| **Additional** | C18 | Malignant neoplasm of colon | 235 | 36 | 271 |
| | C19 | Malignant neoplasm of rectosigmoid junction | 38 | 6 | 44 |
| | C20 | Malignant neoplasm of rectum | 83 | 7 | 90 |
| | C50 | Malignant neoplasm of breast | 500 | 103 | 603 |
| | C61 | Malignant neoplasm of prostate | 501 | 104 | 605 |
| | C85 | Other specified and unspecified types of non-Hodgkin lymphoma | 225 | 59 | 284 |
| | C91 | Lymphoid leukemia | 174 | 35 | 209 |
| | G35 | Multiple sclerosis | 175 | 34 | 209 |
| | K50 | Crohn's disease [regional enteritis] | 275 | 49 | 324 |
| | K51 | Ulcerative colitis | 249 | 53 | 302 |

Event counts indicate patients who developed the corresponding incident endpoint during follow-up in each cohort. Focal endpoints comprised 57 endpoints assigned to the six focal organ systems; 10 additional endpoints were retained to broaden outcome coverage. All 67 endpoints contributed to the unified prior-history and washout exclusions, including those with few or no events in the held-out test set.

**Supplementary Table 3 | Discrimination of five model configurations across 67 individual ICD-10 endpoints in the held-out test set.**

| Organ system | ICD-10 prefix | Condition | Events, n | Demographics | Demographics-laboratory | ECG-only | ECG-demographics | ECG-RISK |
|---|---|---|---|---|---|---|---|---|
| **Heart** | I05 | Rheumatic mitral valve diseases | 23 | 0.706 (0.563-0.829) | 0.686 (0.554-0.811) | 0.770 (0.674-0.869) | 0.786 (0.681-0.893) | 0.811 (0.729-0.891) |
| | I06 | Rheumatic aortic valve diseases | 1 | 0.975 (0.970-0.979) | 0.976 (0.971-0.980) | 0.997 (0.996-0.999) | 0.990 (0.987-0.993) | 0.993 (0.990-0.995) |
| | I07 | Rheumatic tricuspid valve diseases | 100 | 0.779 (0.731-0.818) | 0.809 (0.763-0.848) | 0.861 (0.824-0.892) | 0.872 (0.835-0.901) | 0.871 (0.837-0.900) |
| | I08 | Multiple valve diseases | 165 | 0.793 (0.759-0.828) | 0.820 (0.787-0.849) | 0.854 (0.827-0.878) | 0.856 (0.829-0.882) | 0.868 (0.841-0.891) |
| | I09 | Other rheumatic heart diseases | 14 | 0.822 (0.613-0.942) | 0.814 (0.629-0.933) | 0.806 (0.673-0.922) | 0.882 (0.819-0.939) | 0.873 (0.799-0.936) |
| | I11 | Hypertensive heart disease | 354 | 0.694 (0.668-0.721) | 0.735 (0.708-0.761) | 0.795 (0.771-0.820) | 0.801 (0.778-0.824) | 0.806 (0.783-0.829) |
| | I20 | Angina pectoris | 199 | 0.741 (0.708-0.773) | 0.753 (0.720-0.783) | 0.746 (0.714-0.774) | 0.775 (0.746-0.802) | 0.776 (0.747-0.803) |
| | I21 | Acute myocardial infarction | 327 | 0.730 (0.704-0.755) | 0.761 (0.736-0.785) | 0.769 (0.741-0.792) | 0.774 (0.750-0.797) | 0.787 (0.763-0.809) |
| | I22 | Subsequent myocardial infarction | 4 | 0.165 (0.047-0.431) | 0.226 (0.089-0.452) | 0.353 (0.095-0.995) | 0.188 (0.004-0.771) | 0.300 (0.203-0.881) |
| | I23 | Certain current complications following acute myocardial infarction | 0 | NE | NE | NE | NE | NE |
| | I24 | Other acute ischemic heart diseases | 118 | 0.717 (0.674-0.759) | 0.786 (0.750-0.819) | 0.805 (0.764-0.838) | 0.803 (0.761-0.839) | 0.828 (0.792-0.857) |

| | | | | | | | |
|---|---|---|---|---|---|---|---|
| I25 | Chronic ischemic heart disease | 1,203 | 0.780 (0.766-0.792) | 0.794 (0.782-0.806) | 0.802 (0.790-0.815) | 0.823 (0.811-0.835) | 0.827 (0.815-0.839) |
| I34 | Nonrheumatic mitral valve disorders | 300 | 0.720 (0.693-0.749) | 0.746 (0.719-0.773) | 0.776 (0.748-0.804) | 0.791 (0.763-0.817) | 0.799 (0.774-0.823) |
| I35 | Nonrheumatic aortic valve disorders | 245 | 0.815 (0.785-0.842) | 0.838 (0.811-0.862) | 0.848 (0.824-0.870) | 0.870 (0.849-0.889) | 0.872 (0.850-0.891) |
| I36 | Nonrheumatic tricuspid valve disorders | 26 | 0.640 (0.523-0.760) | 0.709 (0.587-0.816) | 0.730 (0.627-0.822) | 0.717 (0.589-0.848) | 0.734 (0.620-0.845) |
| I37 | Nonrheumatic pulmonary valve disorders | 5 | 0.462 (0.119-0.905) | 0.548 (0.177-0.845) | 0.608 (0.335-0.937) | 0.570 (0.394-0.832) | 0.700 (0.616-0.866) |
| I38 | Endocarditis, valve unspecified | 14 | 0.592 (0.445-0.753) | 0.527 (0.370-0.699) | 0.526 (0.325-0.750) | 0.577 (0.362-0.788) | 0.504 (0.313-0.712) |
| I42 | Cardiomyopathy | 259 | 0.664 (0.628-0.698) | 0.707 (0.674-0.737) | 0.836 (0.810-0.862) | 0.837 (0.811-0.863) | 0.838 (0.813-0.864) |
| I44 | Atrioventricular and left bundle-branch block | 268 | 0.765 (0.736-0.793) | 0.779 (0.751-0.808) | 0.873 (0.848-0.894) | 0.862 (0.835-0.885) | 0.860 (0.832-0.883) |
| I45 | Other conduction disorders | 180 | 0.657 (0.615-0.698) | 0.694 (0.650-0.737) | 0.777 (0.740-0.818) | 0.781 (0.744-0.819) | 0.785 (0.747-0.822) |
| I47 | Paroxysmal tachycardia | 319 | 0.670 (0.639-0.702) | 0.707 (0.678-0.735) | 0.750 (0.716-0.779) | 0.760 (0.729-0.788) | 0.768 (0.739-0.795) |
| I48 | Atrial fibrillation and flutter | 1,042 | 0.782 (0.768-0.794) | 0.795 (0.782-0.808) | 0.830 (0.817-0.843) | 0.839 (0.826-0.850) | 0.841 (0.828-0.852) |
| I50 | Heart failure | 941 | 0.752 (0.737-0.768) | 0.789 (0.775-0.803) | 0.846 (0.833-0.858) | 0.853 (0.841-0.865) | 0.857 (0.846-0.869) |
| I70 | Atherosclerosis | 178 | 0.740 (0.707-0.776) | 0.772 (0.742-0.804) | 0.805 (0.776-0.833) | 0.810 (0.780-0.839) | 0.814 (0.784-0.843) |
| I71 | Aortic aneurysm and dissection | 112 | 0.801 (0.760-0.840) | 0.810 (0.770-0.847) | 0.810 (0.773-0.846) | 0.825 (0.789-0.859) | 0.827 (0.791-0.860) |

| | | | | | | | | |
|---|---|---|---|---|---|---|---|---|
| | I73 | Other peripheral vascular diseases | 267 | 0.722 (0.689-0.754) | 0.738 (0.705-0.767) | 0.747 (0.713-0.778) | 0.767 (0.737-0.795) | 0.765 (0.734-0.794) |
| **Brain** | C71 | Malignant neoplasm of brain | 14 | 0.582 (0.424-0.735) | 0.618 (0.447-0.786) | 0.502 (0.325-0.676) | 0.571 (0.401-0.730) | 0.521 (0.366-0.674) |
| | F01 | Vascular dementia | 54 | 0.890 (0.856-0.917) | 0.903 (0.878-0.927) | 0.823 (0.762-0.876) | 0.896 (0.863-0.923) | 0.898 (0.870-0.924) |
| | F02 | Dementia in other diseases classified elsewhere | 143 | 0.880 (0.851-0.906) | 0.881 (0.852-0.906) | 0.782 (0.749-0.815) | 0.877 (0.849-0.903) | 0.879 (0.852-0.902) |
| | F03 | Unspecified dementia | 285 | 0.901 (0.886-0.915) | 0.900 (0.887-0.914) | 0.819 (0.797-0.842) | 0.903 (0.889-0.917) | 0.899 (0.885-0.913) |
| | G20 | Parkinson's disease | 74 | 0.820 (0.787-0.854) | 0.814 (0.778-0.851) | 0.760 (0.711-0.809) | 0.828 (0.789-0.865) | 0.831 (0.792-0.868) |
| | G30 | Alzheimer's disease | 109 | 0.904 (0.874-0.927) | 0.900 (0.869-0.925) | 0.811 (0.776-0.844) | 0.896 (0.867-0.920) | 0.897 (0.869-0.921) |
| | G81 | Hemiplegia and hemiparesis | 112 | 0.634 (0.583-0.688) | 0.657 (0.605-0.708) | 0.674 (0.621-0.725) | 0.681 (0.627-0.733) | 0.681 (0.631-0.729) |
| | I60 | Nontraumatic subarachnoid hemorrhage | 23 | 0.595 (0.488-0.701) | 0.651 (0.554-0.745) | 0.635 (0.510-0.753) | 0.617 (0.480-0.744) | 0.651 (0.549-0.751) |
| | I61 | Intracerebral haemorrhage | 65 | 0.696 (0.628-0.761) | 0.698 (0.632-0.760) | 0.701 (0.629-0.777) | 0.718 (0.645-0.790) | 0.722 (0.658-0.793) |
| | I62 | Other and unspecified nontraumatic intracranial hemorrhage | 38 | 0.748 (0.679-0.809) | 0.712 (0.645-0.776) | 0.659 (0.584-0.728) | 0.738 (0.667-0.797) | 0.714 (0.642-0.779) |
| | I63 | Cerebral infarction | 227 | 0.716 (0.683-0.748) | 0.725 (0.691-0.756) | 0.744 (0.714-0.774) | 0.753 (0.721-0.782) | 0.753 (0.722-0.783) |
| **Kidney** | C64 | Malignant neoplasm of kidney, except renal pelvis | 61 | 0.752 (0.697-0.809) | 0.766 (0.709-0.819) | 0.704 (0.645-0.763) | 0.746 (0.688-0.804) | 0.764 (0.705-0.816) |

| | | | | | | | | |
|---|---|---|---|---|---|---|---|---|
| | I12 | Hypertensive renal disease | 783 | 0.768 (0.753-0.785) | 0.850 (0.837-0.863) | 0.793 (0.778-0.808) | 0.819 (0.806-0.834) | 0.863 (0.850-0.875) |
| | N17 | Acute kidney failure | 1,374 | 0.696 (0.681-0.711) | 0.750 (0.737-0.763) | 0.717 (0.703-0.731) | 0.737 (0.724-0.750) | 0.767 (0.755-0.780) |
| | N18 | Chronic kidney disease (CKD) | 949 | 0.757 (0.741-0.772) | 0.838 (0.825-0.850) | 0.782 (0.766-0.796) | 0.808 (0.794-0.821) | 0.851 (0.839-0.863) |
| | N19 | Unspecified kidney failure | 11 | 0.718 (0.583-0.856) | 0.728 (0.591-0.868) | 0.635 (0.437-0.827) | 0.726 (0.601-0.843) | 0.732 (0.618-0.852) |
| **Lung** | C34 | Malignant neoplasm of bronchus and lung | 139 | 0.762 (0.730-0.792) | 0.774 (0.743-0.804) | 0.777 (0.741-0.812) | 0.787 (0.756-0.818) | 0.787 (0.753-0.819) |
| | J41 | Simple and mucopurulent chronic bronchitis | 3 | 0.628 (0.380-0.887) | 0.807 (0.572-0.983) | 0.339 (0.215-0.443) | 0.678 (0.454-0.842) | 0.572 (0.301-0.816) |
| | J42 | Unspecified chronic bronchitis | 9 | 0.567 (0.392-0.748) | 0.569 (0.399-0.750) | 0.632 (0.506-0.789) | 0.620 (0.463-0.815) | 0.647 (0.462-0.840) |
| | J43 | Emphysema | 104 | 0.760 (0.718-0.804) | 0.756 (0.713-0.797) | 0.833 (0.798-0.869) | 0.830 (0.795-0.865) | 0.836 (0.798-0.871) |
| | J44 | Other chronic obstructive pulmonary disease | 523 | 0.733 (0.715-0.752) | 0.747 (0.728-0.766) | 0.787 (0.768-0.806) | 0.801 (0.782-0.819) | 0.803 (0.785-0.820) |
| | J47 | Bronchiectasis | 53 | 0.725 (0.654-0.789) | 0.754 (0.696-0.810) | 0.750 (0.670-0.817) | 0.767 (0.703-0.826) | 0.788 (0.726-0.843) |
| | J84 | Other interstitial pulmonary diseases | 69 | 0.696 (0.619-0.762) | 0.712 (0.639-0.771) | 0.725 (0.648-0.789) | 0.729 (0.656-0.794) | 0.736 (0.667-0.797) |
| **Liver** | C22 | Malignant neoplasm of liver and intrahepatic bile ducts | 42 | 0.782 (0.721-0.837) | 0.793 (0.727-0.857) | 0.758 (0.675-0.833) | 0.808 (0.749-0.863) | 0.814 (0.756-0.872) |
| | K70 | Alcoholic liver disease | 91 | 0.740 (0.688-0.790) | 0.849 (0.805-0.884) | 0.768 (0.718-0.816) | 0.819 (0.775-0.859) | 0.873 (0.835-0.905) |

| | | | | | | | | |
|---|---|---|---|---|---|---|---|---|
| | K72 | Hepatic failure, not elsewhere classified | 82 | 0.673 (0.598-0.737) | 0.750 (0.690-0.800) | 0.709 (0.641-0.769) | 0.729 (0.662-0.789) | 0.757 (0.693-0.813) |
| | K74 | Fibrosis and cirrhosis of liver | 144 | 0.679 (0.634-0.720) | 0.747 (0.709-0.787) | 0.700 (0.660-0.741) | 0.736 (0.697-0.776) | 0.775 (0.739-0.808) |
| | K76 | Other diseases of liver | 394 | 0.597 (0.566-0.628) | 0.641 (0.611-0.671) | 0.643 (0.615-0.672) | 0.648 (0.620-0.675) | 0.663 (0.632-0.692) |
| **Pancreas** | C25 | Malignant neoplasm of pancreas | 37 | 0.769 (0.700-0.837) | 0.792 (0.731-0.846) | 0.653 (0.566-0.734) | 0.725 (0.662-0.790) | 0.771 (0.704-0.828) |
| | K85 | Acute pancreatitis | 143 | 0.546 (0.491-0.597) | 0.638 (0.588-0.685) | 0.563 (0.516-0.610) | 0.571 (0.524-0.621) | 0.620 (0.568-0.671) |
| | K86 | Other diseases of pancreas | 118 | 0.619 (0.572-0.666) | 0.678 (0.624-0.725) | 0.648 (0.596-0.698) | 0.652 (0.606-0.699) | 0.673 (0.620-0.720) |
| **Additional** | C18 | Malignant neoplasm of colon | 36 | 0.692 (0.619-0.768) | 0.773 (0.704-0.837) | 0.689 (0.615-0.757) | 0.699 (0.628-0.772) | 0.760 (0.691-0.822) |
| | C19 | Malignant neoplasm of rectosigmoid junction | 6 | 0.596 (0.339-0.847) | 0.793 (0.647-0.901) | 0.702 (0.480-0.845) | 0.674 (0.458-0.853) | 0.761 (0.538-0.898) |
| | C20 | Malignant neoplasm of rectum | 7 | 0.615 (0.436-0.815) | 0.559 (0.323-0.787) | 0.693 (0.509-0.835) | 0.720 (0.492-0.908) | 0.650 (0.414-0.829) |
| | C50 | Malignant neoplasm of breast | 103 | 0.804 (0.769-0.837) | 0.787 (0.755-0.817) | 0.697 (0.653-0.740) | 0.807 (0.772-0.839) | 0.783 (0.750-0.816) |
| | C61 | Malignant neoplasm of prostate | 104 | 0.910 (0.894-0.926) | 0.899 (0.882-0.916) | 0.804 (0.769-0.838) | 0.907 (0.892-0.923) | 0.885 (0.867-0.902) |
| | C85 | Other specified and unspecified types of non-Hodgkin lymphoma | 59 | 0.724 (0.666-0.781) | 0.750 (0.695-0.803) | 0.714 (0.636-0.784) | 0.735 (0.674-0.796) | 0.756 (0.692-0.814) |
| | C91 | Lymphoid leukemia | 35 | 0.766 (0.701-0.829) | 0.811 (0.739-0.873) | 0.599 (0.501-0.697) | 0.758 (0.690-0.821) | 0.800 (0.730-0.857) |
| | G35 | Multiple sclerosis | 34 | 0.665 (0.563-0.765) | 0.691 (0.574-0.799) | 0.540 (0.434-0.634) | 0.650 (0.570-0.731) | 0.639 (0.553-0.727) |

| | | | | | | | |
|---|---|---|---|---|---|---|---|
| K50 | Crohn's disease [regional enteritis] | 49 | 0.709 (0.639-0.777) | 0.705 (0.639-0.774) | 0.450 (0.360-0.559) | 0.588 (0.507-0.662) | 0.499 (0.418-0.575) |
| K51 | Ulcerative colitis | 53 | 0.627 (0.548-0.701) | 0.586 (0.504-0.655) | 0.506 (0.417-0.589) | 0.578 (0.498-0.647) | 0.543 (0.461-0.615) |

Events are counts in the held-out test set. Values are C-indices (95% confidence intervals). Demographics included age, sex and recorded race or ethnicity; Demographics-laboratory additionally included laboratory features; ECG-only used ECG waveforms alone; ECG-demographics additionally included ECG waveforms; and ECG-RISK included ECG waveforms, demographic variables and laboratory features. Confidence intervals were estimated using 1,000 patient-level bootstrap resamples, excluding resamples in which the C-index was not estimable. NE indicates that no C-index could be estimated because no event occurred in the held-out test set. Estimates for low-event endpoints are descriptive and should be interpreted cautiously.

**Supplementary Table 4 | Paired differences in C-index relative to the Demographics model across 67 individual ICD-10 endpoints in the held-out test set.**

| Organ system | ICD-10 prefix | Condition | Events, n | ΔLaboratory | ΔECG | ΔECG-RISK |
|---|---|---|---|---|---|---|
| **Heart** | I05 | Rheumatic mitral valve diseases | 23 | -0.020 (-0.084 to +0.035) | +0.080 (-0.073 to +0.234) | +0.104 (-0.024 to +0.245) |
| | I06 | Rheumatic aortic valve diseases | 1 | +0.001 (-0.003 to +0.006) | +0.015 (+0.010 to +0.021) | +0.018 (+0.012 to +0.023) |
| | I07 | Rheumatic tricuspid valve diseases | 100 | +0.030 (+0.006 to +0.055) | +0.093 (+0.060 to +0.127) | +0.093 (+0.062 to +0.124) |
| | I08 | Multiple valve diseases | 165 | +0.027 (+0.004 to +0.051) | +0.063 (+0.037 to +0.092) | +0.074 (+0.048 to +0.104) |
| | I09 | Other rheumatic heart diseases | 14 | -0.009 (-0.074 to +0.044) | +0.060 (-0.086 to +0.256) | +0.051 (-0.099 to +0.250) |
| | I11 | Hypertensive heart disease | 354 | +0.040 (+0.022 to +0.059) | +0.106 (+0.081 to +0.131) | +0.112 (+0.089 to +0.134) |
| | I20 | Angina pectoris | 199 | +0.011 (-0.004 to +0.028) | +0.033 (+0.016 to +0.053) | +0.035 (+0.015 to +0.057) |
| | I21 | Acute myocardial infarction | 327 | +0.031 (+0.017 to +0.047) | +0.045 (+0.028 to +0.062) | +0.057 (+0.038 to +0.076) |
| | I22 | Subsequent myocardial infarction | 4 | +0.061 (+0.030 to +0.128) | +0.023 (-0.134 to +0.222) | +0.136 (+0.039 to +0.283) |
| | I23 | Certain current complications following acute myocardial infarction | 0 | NE | NE | NE |
| | I24 | Other acute ischemic heart diseases | 118 | +0.069 (+0.039 to +0.100) | +0.086 (+0.052 to +0.118) | +0.111 (+0.076 to +0.144) |
| | I25 | Chronic ischemic heart disease | 1,203 | +0.014 (+0.009 to +0.020) | +0.043 (+0.035 to +0.051) | +0.047 (+0.039 to +0.056) |
| | I34 | Nonrheumatic mitral valve disorders | 300 | +0.026 (+0.010 to +0.043) | +0.071 (+0.047 to +0.094) | +0.078 (+0.053 to +0.102) |
| | I35 | Nonrheumatic aortic valve disorders | 245 | +0.023 (+0.011 to +0.036) | +0.055 (+0.036 to +0.073) | +0.057 (+0.036 to +0.078) |
| | I36 | Nonrheumatic tricuspid valve disorders | 26 | +0.069 (+0.011 to +0.127) | +0.077 (-0.007 to +0.165) | +0.094 (+0.007 to +0.178) |
| | I37 | Nonrheumatic pulmonary valve disorders | 5 | +0.086 (-0.058 to +0.174) | +0.108 (-0.195 to +0.549) | +0.238 (-0.030 to +0.579) |
| | I38 | Endocarditis, valve unspecified | 14 | -0.065 (-0.157 to +0.051) | -0.016 (-0.223 to +0.229) | -0.088 (-0.305 to +0.178) |
| | I42 | Cardiomyopathy | 259 | +0.043 (+0.021 to +0.065) | +0.174 (+0.138 to +0.206) | +0.175 (+0.139 to +0.208) |
| | I44 | Atrioventricular and left bundle-branch block | 268 | +0.015 (+0.003 to +0.026) | +0.098 (+0.073 to +0.123) | +0.096 (+0.070 to +0.122) |

| | | | | | | |
|---|---|---|---|---|---|---|
| | I45 | Other conduction disorders | 180 | +0.037 (+0.007 to +0.067) | +0.124 (+0.082 to +0.165) | +0.128 (+0.089 to +0.168) |
| | I47 | Paroxysmal tachycardia | 319 | +0.037 (+0.021 to +0.052) | +0.089 (+0.064 to +0.116) | +0.098 (+0.072 to +0.124) |
| | I48 | Atrial fibrillation and flutter | 1,042 | +0.014 (+0.009 to +0.020) | +0.057 (+0.048 to +0.066) | +0.059 (+0.051 to +0.068) |
| | I50 | Heart failure | 941 | +0.037 (+0.028 to +0.045) | +0.101 (+0.088 to +0.113) | +0.105 (+0.092 to +0.117) |
| | I70 | Atherosclerosis | 178 | +0.032 (+0.007 to +0.057) | +0.070 (+0.044 to +0.095) | +0.074 (+0.046 to +0.101) |
| | I71 | Aortic aneurysm and dissection | 112 | +0.009 (-0.004 to +0.022) | +0.024 (+0.003 to +0.046) | +0.026 (+0.003 to +0.048) |
| | I73 | Other peripheral vascular diseases | 267 | +0.015 (-0.001 to +0.033) | +0.045 (+0.027 to +0.066) | +0.043 (+0.022 to +0.066) |
| **Brain** | C71 | Malignant neoplasm of brain | 14 | +0.036 (-0.065 to +0.138) | -0.012 (-0.099 to +0.081) | -0.062 (-0.141 to +0.020) |
| | F01 | Vascular dementia | 54 | +0.013 (-0.006 to +0.034) | +0.006 (-0.005 to +0.018) | +0.009 (-0.008 to +0.027) |
| | F02 | Dementia in other diseases classified elsewhere | 143 | +0.001 (-0.009 to +0.010) | -0.003 (-0.010 to +0.004) | -0.001 (-0.013 to +0.011) |
| | F03 | Unspecified dementia | 285 | 0.000 (-0.008 to +0.007) | +0.002 (-0.001 to +0.006) | -0.002 (-0.010 to +0.006) |
| | G20 | Parkinson's disease | 74 | -0.006 (-0.018 to +0.006) | +0.009 (-0.007 to +0.024) | +0.011 (-0.008 to +0.031) |
| | G30 | Alzheimer's disease | 109 | -0.004 (-0.014 to +0.007) | -0.007 (-0.014 to 0.000) | -0.006 (-0.017 to +0.005) |
| | G81 | Hemiplegia and hemiparesis | 112 | +0.023 (-0.008 to +0.054) | +0.047 (+0.008 to +0.092) | +0.047 (+0.014 to +0.087) |
| | I60 | Nontraumatic subarachnoid hemorrhage | 23 | +0.056 (-0.006 to +0.125) | +0.022 (-0.059 to +0.112) | +0.056 (-0.011 to +0.128) |
| | I61 | Intracerebral haemorrhage | 65 | +0.001 (-0.021 to +0.026) | +0.022 (-0.015 to +0.062) | +0.026 (-0.008 to +0.063) |
| | I62 | Other and unspecified nontraumatic intracranial hemorrhage | 38 | -0.035 (-0.071 to +0.004) | -0.010 (-0.040 to +0.021) | -0.034 (-0.066 to +0.002) |
| | I63 | Cerebral infarction | 227 | +0.008 (-0.004 to +0.023) | +0.037 (+0.018 to +0.057) | +0.037 (+0.021 to +0.057) |
| **Kidney** | C64 | Malignant neoplasm of kidney, except renal pelvis | 61 | +0.014 (-0.020 to +0.053) | -0.006 (-0.031 to +0.021) | +0.012 (-0.023 to +0.050) |
| | I12 | Hypertensive renal disease | 783 | +0.082 (+0.069 to +0.096) | +0.051 (+0.040 to +0.063) | +0.095 (+0.080 to +0.110) |
| | N17 | Acute kidney failure | 1,374 | +0.054 (+0.043 to +0.065) | +0.041 (+0.031 to +0.052) | +0.071 (+0.059 to +0.083) |
| | N18 | Chronic kidney disease (CKD) | 949 | +0.081 (+0.069 to +0.095) | +0.051 (+0.040 to +0.063) | +0.094 (+0.081 to +0.108) |
| | N19 | Unspecified kidney failure | 11 | +0.009 (-0.053 to +0.075) | +0.008 (-0.071 to +0.085) | +0.013 (-0.063 to +0.081) |

| | | | | | | |
|---|---|---|---|---|---|---|
| **Lung** | C34 | Malignant neoplasm of bronchus and lung | 139 | +0.013 (-0.006 to +0.030) | +0.026 (-0.003 to +0.053) | +0.025 (-0.005 to +0.053) |
| | J41 | Simple and mucopurulent chronic bronchitis | 3 | +0.179 (-0.310 to +0.602) | +0.050 (-0.045 to +0.097) | -0.056 (-0.348 to +0.435) |
| | J42 | Unspecified chronic bronchitis | 9 | +0.002 (-0.032 to +0.043) | +0.052 (-0.070 to +0.219) | +0.080 (-0.010 to +0.228) |
| | J43 | Emphysema | 104 | -0.004 (-0.038 to +0.028) | +0.070 (+0.028 to +0.113) | +0.075 (+0.027 to +0.123) |
| | J44 | Other chronic obstructive pulmonary disease | 523 | +0.013 (+0.001 to +0.025) | +0.067 (+0.050 to +0.084) | +0.069 (+0.053 to +0.086) |
| | J47 | Bronchiectasis | 53 | +0.028 (-0.010 to +0.073) | +0.042 (-0.005 to +0.087) | +0.062 (+0.004 to +0.130) |
| | J84 | Other interstitial pulmonary diseases | 69 | +0.016 (-0.016 to +0.054) | +0.033 (-0.004 to +0.073) | +0.040 (+0.005 to +0.079) |
| **Liver** | C22 | Malignant neoplasm of liver and intrahepatic bile ducts | 42 | +0.012 (-0.034 to +0.060) | +0.026 (-0.019 to +0.072) | +0.032 (-0.014 to +0.084) |
| | K70 | Alcoholic liver disease | 91 | +0.109 (+0.065 to +0.162) | +0.079 (+0.045 to +0.115) | +0.133 (+0.087 to +0.186) |
| | K72 | Hepatic failure, not elsewhere classified | 82 | +0.076 (+0.023 to +0.128) | +0.056 (+0.013 to +0.099) | +0.084 (+0.026 to +0.143) |
| | K74 | Fibrosis and cirrhosis of liver | 144 | +0.069 (+0.029 to +0.104) | +0.057 (+0.027 to +0.086) | +0.096 (+0.058 to +0.138) |
| | K76 | Other diseases of liver | 394 | +0.044 (+0.018 to +0.071) | +0.051 (+0.021 to +0.083) | +0.066 (+0.036 to +0.096) |
| **Pancreas** | C25 | Malignant neoplasm of pancreas | 37 | +0.023 (-0.018 to +0.068) | -0.044 (-0.077 to -0.012) | +0.002 (-0.048 to +0.055) |
| | K85 | Acute pancreatitis | 143 | +0.091 (+0.038 to +0.146) | +0.025 (-0.033 to +0.080) | +0.074 (+0.009 to +0.138) |
| | K86 | Other diseases of pancreas | 118 | +0.058 (+0.013 to +0.107) | +0.033 (-0.013 to +0.085) | +0.053 (+0.002 to +0.108) |
| **Additional** | C18 | Malignant neoplasm of colon | 36 | +0.081 (+0.007 to +0.169) | +0.007 (-0.015 to +0.028) | +0.068 (-0.007 to +0.147) |
| | C19 | Malignant neoplasm of rectosigmoid junction | 6 | +0.197 (+0.043 to +0.406) | +0.078 (-0.034 to +0.184) | +0.165 (-0.009 to +0.332) |
| | C20 | Malignant neoplasm of rectum | 7 | -0.056 (-0.276 to +0.219) | +0.105 (+0.008 to +0.204) | +0.035 (-0.097 to +0.168) |
| | C50 | Malignant neoplasm of breast | 103 | -0.017 (-0.046 to +0.012) | +0.003 (-0.016 to +0.021) | -0.021 (-0.057 to +0.013) |
| | C61 | Malignant neoplasm of prostate | 104 | -0.011 (-0.021 to -0.001) | -0.002 (-0.008 to +0.003) | -0.024 (-0.040 to -0.010) |
| | C85 | Other specified and unspecified types of non-Hodgkin lymphoma | 59 | +0.026 (-0.013 to +0.063) | +0.011 (-0.017 to +0.040) | +0.032 (-0.013 to +0.075) |
| | C91 | Lymphoid leukemia | 35 | +0.045 (+0.012 to +0.076) | -0.007 (-0.030 to +0.015) | +0.034 (-0.005 to +0.069) |
| | G35 | Multiple sclerosis | 34 | +0.026 (-0.020 to +0.076) | -0.015 (-0.058 to +0.022) | -0.026 (-0.102 to +0.044) |
| | K50 | Crohn's disease [regional enteritis] | 49 | -0.004 (-0.081 to +0.078) | -0.121 (-0.205 to -0.036) | -0.210 (-0.312 to -0.118) |

| | | | | | |
|---|---|---|---|---|---|
| K51 | Ulcerative colitis | 53 | -0.041 (-0.090 to +0.009) | -0.050 (-0.112 to +0.015) | -0.085 (-0.160 to -0.011) |

Events are counts in the held-out test set. Values are paired differences in C-index (95% confidence intervals) estimated using 1,000 paired patient-level bootstrap resamples. ΔLaboratory denotes Demographics-laboratory minus Demographics, ΔECG denotes ECG-demographics minus Demographics and ΔECG-RISK denotes ECG-RISK minus Demographics. Positive values favor the augmented model. NE indicates that the difference was not estimable because no event occurred in the held-out test set. Estimates for low-event endpoints are descriptive.

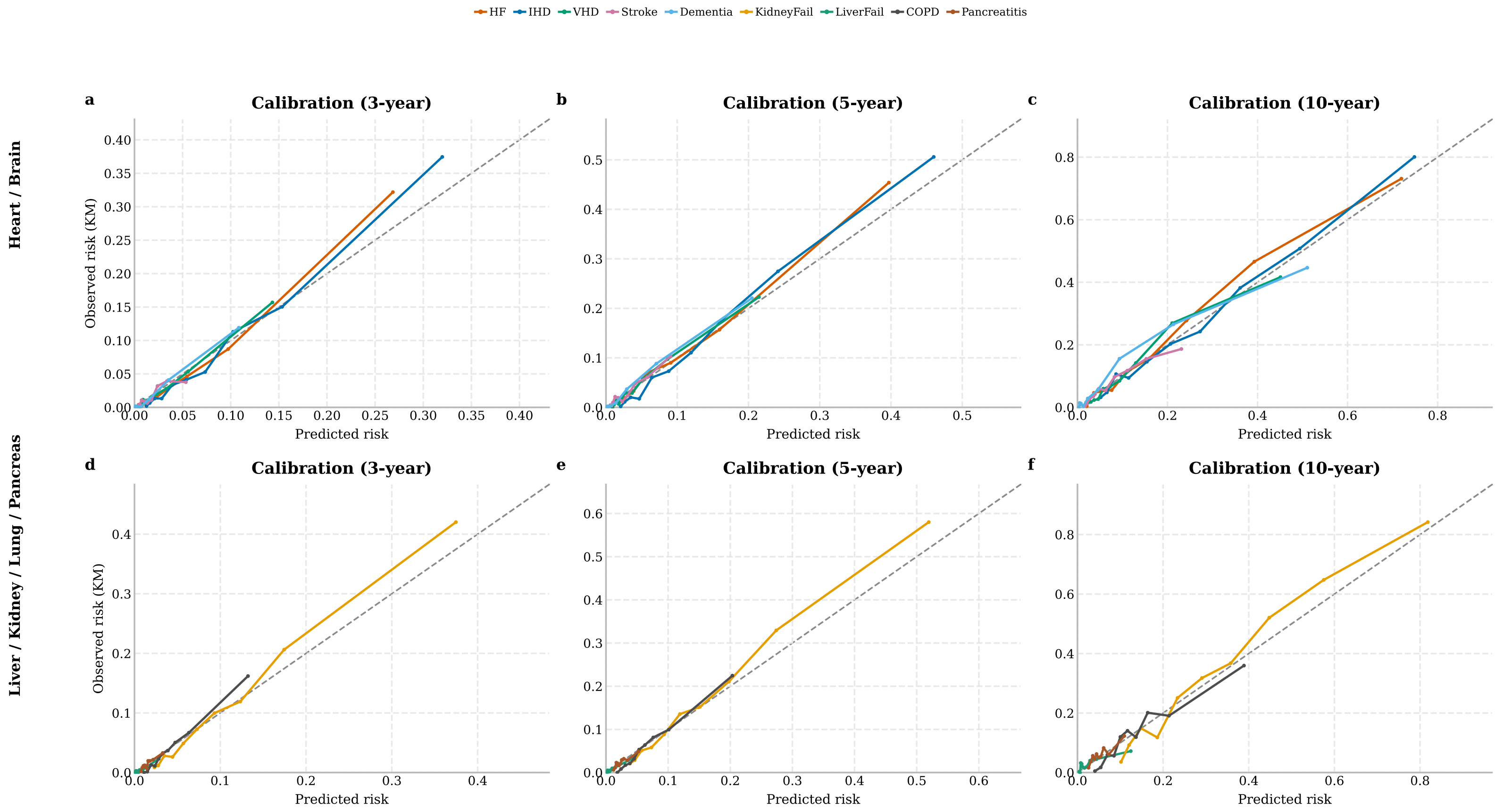


**Supplementary Fig. 1 | Calibration of ECG-RISK for representative aggregated clinical outcomes.**

Calibration in the held-out test set is shown at 3 years (**a,d**), 5 years (**b,e**) and 10 years (**c,f**). The top row presents aggregated heart and brain outcomes, and the bottom row presents liver, kidney, lung and pancreatic outcomes. Patients were grouped into deciles of predicted risk. Points indicate mean predicted risk and Kaplan–Meier estimates of observed risk within each retained bin; lines connect adjacent bins, and diagonal dashed lines indicate perfect calibration. Bins containing fewer than 200 patients or fewer than 10 patients at risk at the respective horizon were omitted. Predicted risks were obtained using fold-specific Cox recalibration models fitted to out-of-fold development predictions and applied to held-out test-set scores.

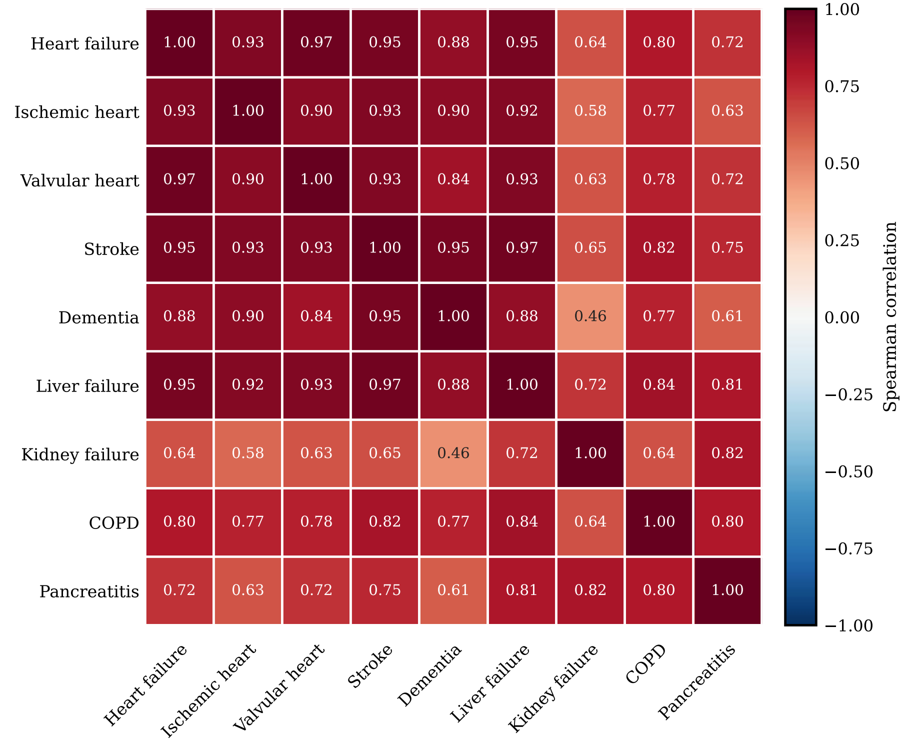


**Supplementary Fig. 2 | Correlations among ECG-RISK scores for representative aggregated clinical outcomes.**

The heatmap shows pairwise Spearman correlations between outcome-specific ECG-RISK log-risk scores for nine representative aggregated clinical outcomes in the held-out test set. Cell values indicate Spearman correlation coefficients. These correlations quantify shared variation among model-derived scores and should not be interpreted as evidence of disease co-occurrence, biological dependence or causal relationships.

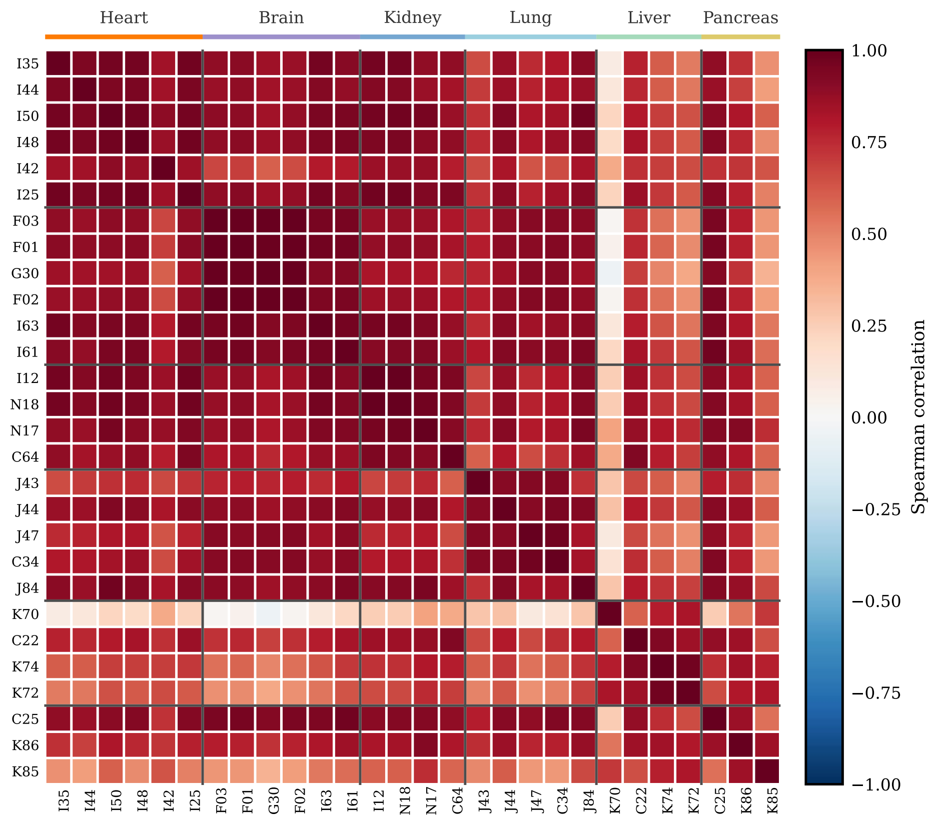


**Supplementary Fig. 3 | Correlations among ECG-RISK scores for representative individual ICD-10 endpoints.**

The heatmap shows pairwise Spearman correlations between endpoint-specific ECG-RISK log-risk scores for the 28 individual ICD-10 endpoints reported in main-text Table 2, calculated across patients in the held-out test set. Endpoints are grouped by organ system and ordered consistently with Table 2. Cell values indicate Spearman correlation coefficients. These correlations characterize similarity among model outputs rather than associations between observed disease occurrences.
